# Serum lipidomics identifies outcome signatures in patients with traumatic brain injury, presenting with Glasgow Coma Scale score of 13-15

**DOI:** 10.64898/2026.09.07.26362408

**Authors:** Henrique C. Ribeiro, Alex M. Dickens, Daniel Duberg, Jussi P. Posti, Andrew I. R. Maas, András Büki, Tuulia Hyötyläinen, David K. Menon, Matej Orešič, CENTER-TBI Participants and Investigators

**Author notes:** A full list of members and their affiliations appear at the end of the manuscript, at Appendix 1.

## Abstract

Patients presenting with a Glasgow Coma Scale (GCS) score of 13–15, conventionally labelled mild traumatic brain injury (mTBI), account for most hospital-presenting TBI; a substantial proportion have not recovered to pre-injury health at six months, fewer than one in ten receives structured follow-up, and at presentation they are largely indistinguishable from those who recover. Prognostic models based on clinical variables discriminate poorly here, and established protein biomarkers add little. Lipidomics reflects membrane turnover and repair rather than injury magnitude, offering a potentially complementary perspective.

Blood collected within 24 h of injury from 929 patients with mTBI in the Collaborative European NeuroTrauma Effectiveness Research in Traumatic Brain Injury (CENTER-TBI) cohort was processed to serum and analysed by Ultra-high performance liquid chromatography – Quadrupole-Time-of-flight mass spectrometry (UHPLC-QTOF). After quality-control, 278 lipid features were retained and clustered by Gaussian mixture modelling. Associations between lipid features, six protein biomarkers, Corticosteroid Randomisation After Significant Head injury (CRASH) clinical variables and 6-month Glasgow Outcome Scale–Extended (GOSE) outcome were assessed by Spearman correlation and Jonckheere–Terpstra trend tests. Variables were selected by ordinal elastic-net and modelled by proportional odds logistic regression, comparing CRASH-only with CRASH-plus-marker models, built in a development set (n = 411 with complete covariates) and assessed in a test set (n = 367).

Lipid clusters rich in lysophosphatidylcholines (LPCs), sphingomyelins/ceramides and ether-linked phosphatidylcholines (PC-O) correlated positively with favourable outcome and inversely with the injury proteins. Summed choline-containing lipid intensity rose across GOSE (p < 0.0001). Elastic-net selected three proteins (GFAP, NfL, S100B) and seven lipid features, three annotated (LPC(18:2), LPC(20:5), PC(O-34:3)). Adding lipids and proteins increased the area under the receiver operating characteristic curve (AUC) at every threshold, reaching nominal significance at GOSE ≥ 7 (AUC 0.728, 95% CI 0.671–0.785, p = 0.022), not surviving correction for four comparisons. The averaged c-index rose from 0.720 to 0.751 and Nagelkerke R^2^ from 0.125 to 0.242. The three proteins and three annotated lipids showed opposing trends across GOSE (all p < 0.001).

The acute serum lipidome is associated with 6-month functional outcome after mTBI. LPCs and PC-Os moved opposite to the injury proteins and may index membrane repair rather than injury burden, offering pathophysiological insight and possible therapeutic targets. Adding them to clinical variables gave modest gains, greatest at the upper end of the outcome scale, where existing models perform worse. The gain is small and exploratory; replication with targeted assays is needed first.

## Introduction

Traumatic brain injury (TBI) has the highest incidence of all neurological disorders, and it is recognised as a large public health and societal problem.^1,2^ TBI is characterised not only as an acute disorder, but as a multi-faceted and dynamic chronic neurological condition triggered by a primary mechanical injury to the brain, leading to a series of secondary events.^1,3^ Patients with TBI show substantial variation in clinical severity, best characterised by emerging multivariable frameworks, such as the CBI-M framework.^4^ However TBI has been canonically classified according to the Glasgow Coma Scale sum score (GCS) into mild (mTBI; GCS 13–15), moderate (moTBI; GCS 9–12) or severe (sTBI; GCS < 9).^5^ The Glasgow Outcome Scale–Extended (GOSE) classifies the functional outcome of an individual patient after TBI into eight categories, ranging from 1 (death) to 8 (upper good recovery).^6^ Characterising each patient accurately in the acute phase and at later time points is important to provide insight into recovery trajectories, and to identify individuals at higher risk of a suboptimal outcome with an ultimate goal to facilitate timely allocation of healthcare resources to the patients who most require them.^7^ In everyday practice, however, that allocation still rests almost entirely on clinical judgement at the bedside, and it is at the mild end of the severity range that this judgement is least reliable.

Although mild TBI (mTBI) is conventionally defined by a GCS score of 13–15, consensus criteria now place greater weight on other clinical features than the GCS alone.^8^ Despite the label, around 50% of patients with mTBI presenting to hospital have not returned to their pre-injury level of health six months after injury, a finding replicated on both sides of the Atlantic,^9,10^ and fewer than 10% of patients discharged after presenting to a European emergency department receive any structured follow-up.^1^

This leads to an unmet need: a test, available at or soon after presentation, that identifies apparently mild-injury patients at genuine risk of incomplete recovery who would benefit from follow-up. Prognostic models addressing this need have been studied extensively across cohorts in recent years, aiming to improve prediction accuracy using clinical variables and blood biomarkers such as proteins and lipids. Proteins such as neurofilament light (NfL), S100 calcium-binding protein B (S100B), ubiquitin C-terminal hydrolase L1 (UCH-L1), and glial fibrillary acidic protein (GFAP) have been evaluated for their viability as injury markers after acute TBI,^3,11–15^ with primary focus on moTBI and sTBI subsets. Where their incremental value has been formally quantified, the gains have been real but modest: adding six serum proteins to the IMPACT and CRASH models raised explained variance significantly, although by less than 5% in absolute terms, while changing the concordance statistic (c-index) by only about 0.01.^16^ Blood-based markers have nonetheless already entered clinical practice in mTBI, where GFAP and UCH-L1 are used to rule out intracranial injury and so avoid unnecessary CT imaging;^17^ combining biomarkers with imaging measures further improves prediction of recovery in patients with mTBI and a normal CT.^18^ These precedents show that a blood test can change management in mTBI when the clinical question is narrowly and correctly framed. Proteins, however, cannot explain metabolic alterations and repair mechanisms, which can be better assessed by analysing the brain lipids, which represent more than 60% of the brain dry weight.^19^ The comprehensive study of lipids in a cell, organ or biological system is called lipidomics,^20^ and it can be strongly appreciated in the context of TBI since lipid alterations occur in both brain and blood after injury and respond very quickly to external stimuli, such as injuries and trauma. This is advantageous for diagnostic and prognostic purposes,^21^ and to evaluate alterations in energy metabolism, membrane degradation, inflammatory cascade and oxidative damage,^19^ which are known processes that take place after acute TBI. We previously reported that the serum lipidome associates with the full spectrum of GCS and of patient outcomes.^22^

A further consideration when building prognostic models for TBI is how the model itself should be constructed. One of the main scales used for outcome assessment is the GOSE scale, which is itself an ordinal scale.^6^ One common strategy for analysing TBI data is to dichotomise the outcome scale,^23^ thereby discarding the information contained in the rank ordering of the outcome scale. An alternative is to train ordinal outcome prediction models that return probabilities at each GOSE threshold within a single model spanning the entire outcome scale,^24^ allowing users to interpret the probability of different levels of recovery, giving a more realistic and honest output of the prediction model.

Here we ask whether the acute circulating lipidome carries prognostic information in mTBI that clinical variables and established injury proteins do not. We hypothesised that, because circulating lipids reflect membrane turnover and repair rather than the magnitude of the initial mechanical insult, they would be complementary to, rather than redundant with, astroglial and axonal injury proteins, and that any added value would be greatest at the upper end of the outcome scale, where clinical models discriminate least well. To test this, we characterised the serum lipidome of patients with mTBI sampled within 24 h of injury in a subset of individuals from the Collaborative European NeuroTrauma Effectiveness Research in Traumatic Brain Injury (CENTER-TBI) cohort, and examined the correlations between lipid clusters, Corticosteroid Randomisation After Significant Head injury (CRASH) prognostic variables,^25^ and protein biomarkers available in the CENTER-TBI database. We then defined a marker panel comprising proteins and lipids and built an ordinal prediction model of 6-month outcome, with CRASH clinical variables as covariates, in order to quantify the incremental value of the selected lipids and proteins over a covariate-only model across the whole GOSE scale rather than at a single dichotomised cut-point. Finally, we examined trends related to injury severity for the selected lipids and for the summed intensity of all choline-containing lipids.

## Materials and methods

### Participants

The CENTER-TBI study recruited 4509 patients from 18 European countries and Israel (https://www.center-tbi.eu/, registered at clinicaltrials. gov NCT02210221),^2^ containing data from 65 centres, collected between Dec 19, 2014, and Dec 17, 2017. Clinical data was accessed using the Mica/Opal platform, core version 3.1.

The data collected under the CENTER-TBI framework contains information regarding the severity of the patients’ injury, based on GCS, and the level of intervention of their treatment, based on the admission status.

Blood samples of the patients were collected within 24 h of injury into gel-separator tubes for serum and centrifuged within 60 min. Serum was then processed, aliquoted (8 x 0.5 mL) and stored at −80 °C locally and shipped on dry ice to the CENTER-TBI central biobank (Pécs, Hungary) for storage.

Samples were divided as development and test set. Both were drawn from the same CENTER-TBI biobank, sampled and stored under a single protocol and analysed in the same laboratory on the same instrument between 2020 and 2022. The development set contains patients with moderate and severe as well as mild TBI; only its mTBI subset is used here. The held-out test set was assembled as an mTBI subcohort from the outset. Part of the data from the development set was reported previously, although our previous work was focused on patients across the full spectrum of TBI.^22^ The initial set comprised 2143 samples (1584 in the development set and 559 in the test set). All patients signed informed consent and ethical approvals, which is described in detail in the Supplementary material.

### Lipidomics analysis

The lipidomic platform used in this study is described in detail elsewhere.^22^ In short, a modified version of the Folch procedure was used in the analysis.^26^ The relevant internal standards, calibration curves, instrument descriptions and UHPLC-MS sample analysis for this study are described in the Supplementary Material and are also described in detail by Thomas *et al.*.^22^

### Study design

After batch correction, patients were selected against pre-specified criteria: an admission GCS score of 13–15 (*i.e.*, mTBI only), age 18 years or older, consent to participate not withdrawn, and an available GOSE assessment at 6 months after injury. The CRASH prognostic model draws on a separate variable, the Glasgow Coma Scale sum score at admission, which was not recorded for 119 of the included patients (**Table 1**); these patients therefore met the mTBI inclusion criterion but could not enter the ordinal models, which required complete CRASH covariates. The resulting analysis set contained 929 patients (503 in the development set and 426 in the held-out test set) and 278 lipids, containing several different lipid classes, such as phosphatidylcholines and ether-linked phosphatidylcholines (PC; O-PC), lysophosphatidylcholines (LPC), phosphatidylethanolamines and plasmalogens phosphatidylethanolamines (PE, P-PE), phosphatidylinositols (PI), phosphatidylglycerols (PG), phosphatidylserines (PS), sphingomyelins (SM), triacylglycerols (TG), ceramides (Cer) and cholesterol esters (CE). A schematic with the study design is presented as **Figure 1**.

**Figure 1.**
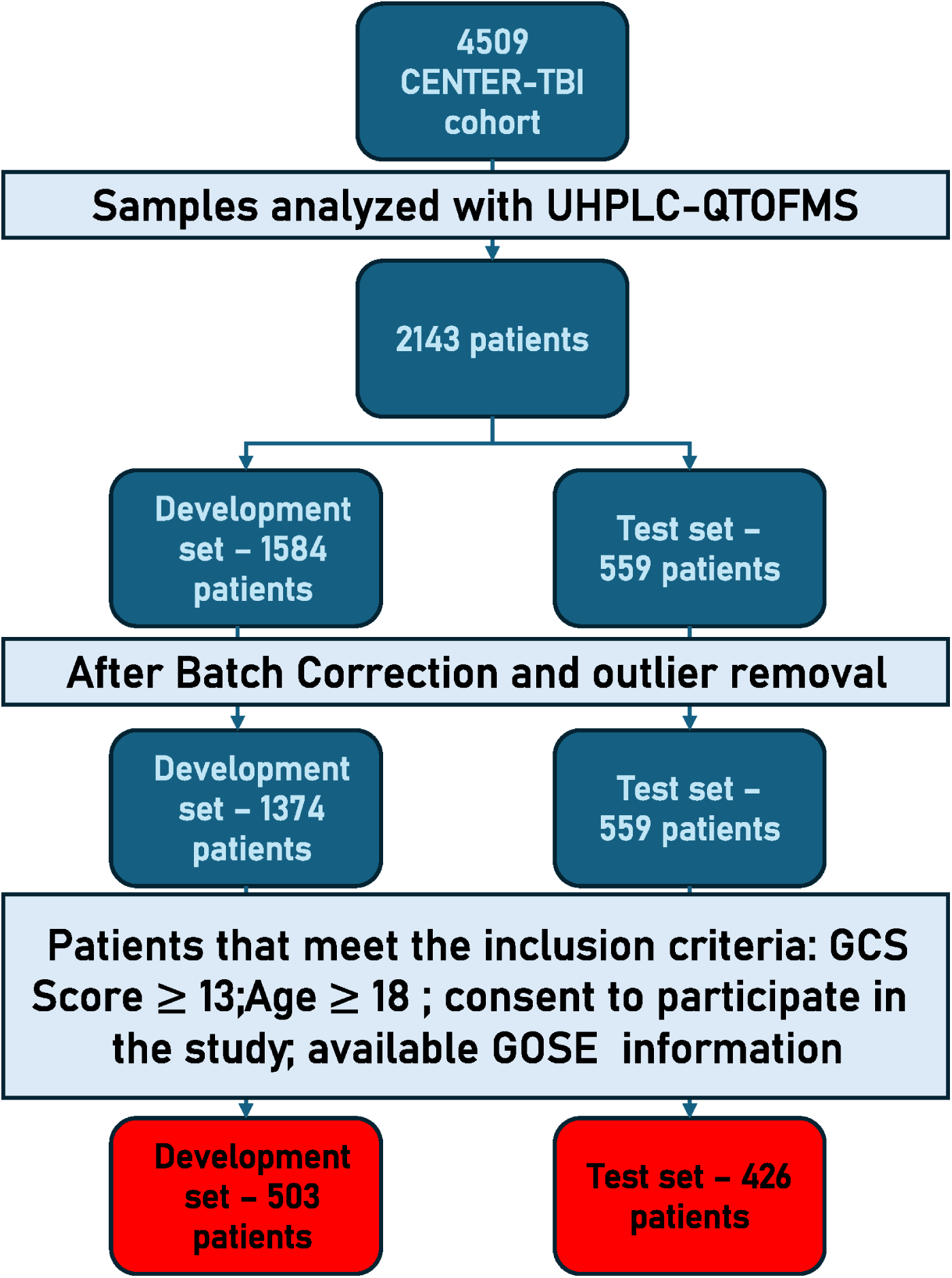
Study design. CENTER-TBI samples were split as a development set and a test set and subjected to batch correction and outlier removal. Afterwards, they were selected according to the following inclusion criteria: GCS score of 13–15, age 18 years or older, consent to participate not withdrawn, and an available GOSE assessment 6 months after injury. Severity classification for this criterion used the injury-severity GCS recorded in the CENTER-TBI core dataset.

**Table 1.**
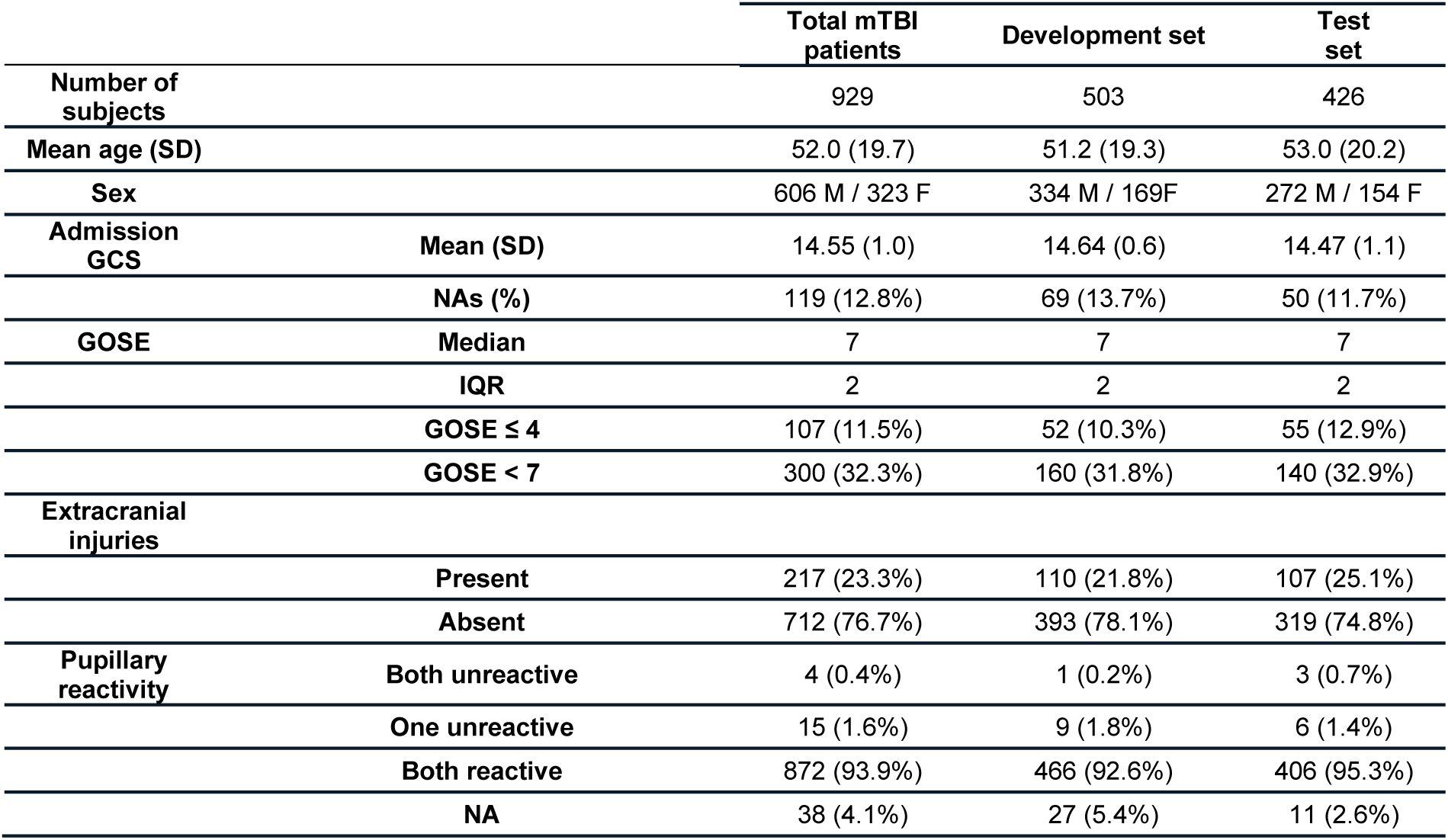
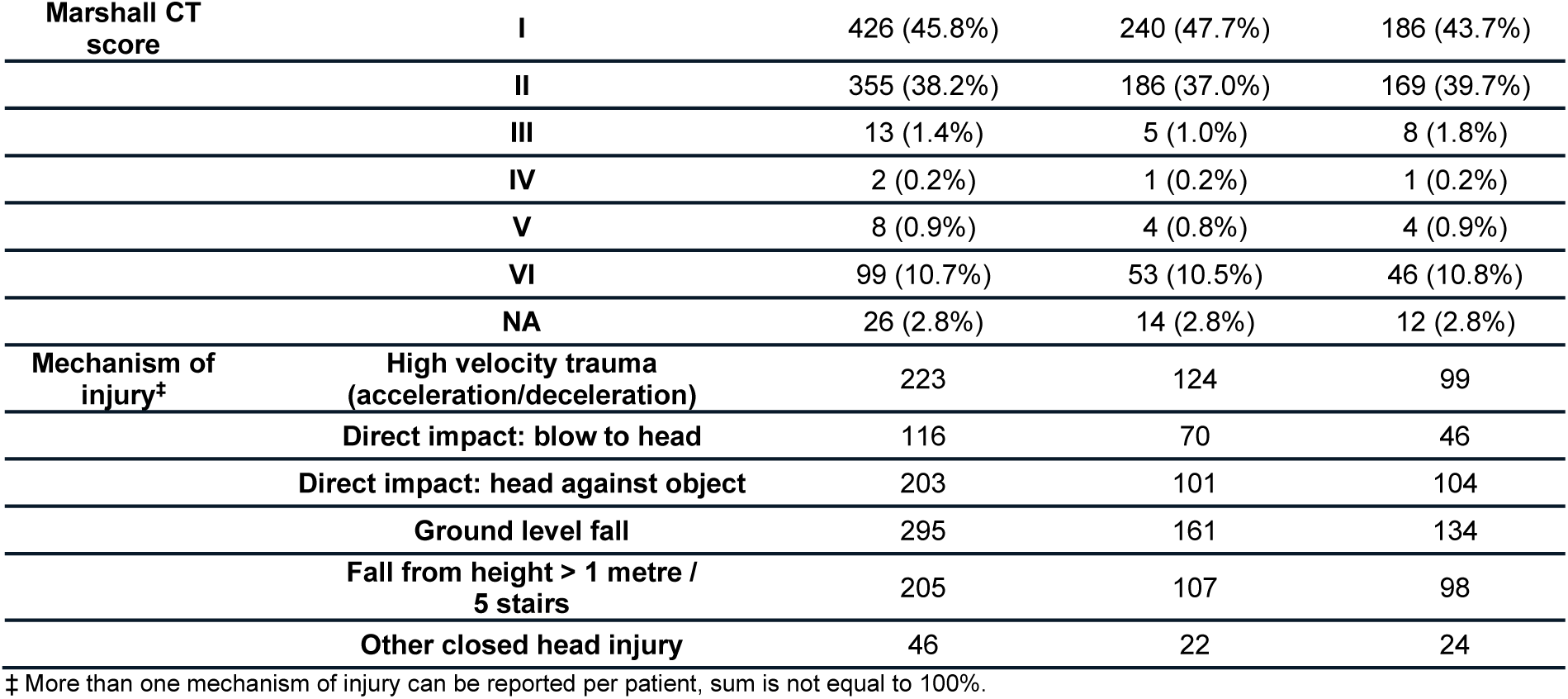
Clinical and demographic data for the selected mTBI patients in CENTER-TBI cohort.

Because the ordinal models required complete data for all CRASH covariates, a further 92 patients in the development set and 59 in test set were excluded at the modelling stage - reflecting 119 patients with no recorded admission GCS and 38 with no recorded pupillary reactivity, of whom 6 were missing both - leaving 411 patients for model development and 367 for held-out assessment; the correlation, choline and trend analyses used all 929 patients.

### Statistical analysis

All analyses were performed in R (v4.4.3).^27^ Preprocessing (imputation, transformation, scaling) was fitted on the development set and applied unchanged to the test set to prevent leakage; batch correction used QC-based correction within each analytical batch. Missing lipid values were imputed with half the minimum observed value per feature, followed by log-transformation and autoscaling.

Unsupervised lipid clustering was performed using model-based Gaussian mixture modelling (*mclust*), ^28^ with clusters selected by BIC and cluster scores computed as mean lipid abundance. Spearman correlations (*Hmisc*) between lipid clusters, protein biomarkers (S100B, NSE, GFAP, UCH-L1, NfL, T-Tau), GOSE outcome, and CRASH covariates^25^ were FDR-corrected (Benjamini-Hochberg) and visualised as a heatmap (*pheatmap*). Choline-containing lipid intensities (PCs, LPCs, SMs) were summed and tested for a monotonic trend across GOSE levels using the Jonckheere-Terpstra test^29,30^ (*DescTools*).

To evaluate a lipid signature’s predictive value, GOSE was collapsed into five ordered levels and modelled with proportional-odds ordinal logistic regression (*MASS::polr*). Predictor selection used ordinal elastic-net (*ordinalNet*, α = 0.5)^31^ with a 20-run stability selection scheme (80% subsampling, 5-fold CV; variables retained if selected in >70% of runs) on a complete-case development set (n = 411). Two models (CRASH covariates alone, and CRASH covariates plus selected lipids and proteins) were compared on a held-out test set (n = 367) using DeLong-tested AUCs (*pROC*, bootstrap-smoothed ROC, 2000 iterations) at each cumulative GOSE threshold, the average dichotomous c-index, and pseudo-Nagelkerke R² (CRASH plus proteins-only and CRASH plus lipids-only variants were also assessed as a supplementary analysis). Internal validation used bootstrap optimism-correction (2000 iterations). Given four threshold comparisons, both unadjusted and Benjamini-Hochberg-corrected p-values are reported, with threshold-specific results treated as exploratory. A multiple-imputation sensitivity analysis (*mice*, 30 datasets, Rubin’s rules) assessed robustness to missing clinical covariates. Full methodological detail is provided in the Supplementary Material.

## Results

### Study setting

In this study we analysed samples from the CENTER-TBI cohort using a UHPLC-MS-based lipidomics platform, acquired in Örebro University, Sweden. The development set comprised 503 patients and the held-out test set 426 patients. After exclusion of patients with incomplete CRASH covariates, data from 411 and 367 patients respectively, were entered into the ordinal models.

Clinical and demographic characteristics of the 929 included patients are shown overall and by set in **Table 1**.

### Lipid cluster correlations with clinical covariates and proteins

The 278 lipid features resolved into 9 lipid clusters (LC) by Gaussian mixture modelling in the mclust algorithm.^28^ The full list of all lipid variables, their cluster assignments, *m/z*, retention time, adducts and MSI identification level^32^ is presented on **Table S1**. In all 929 patients (development and test sets combined), LC 2, 3 and 4 correlated negatively with five of the six protein biomarkers (GFAP, UCH-L1, T-Tau, S100B and NfL), weaklier with NSE, and positively with 6-month GOSE (**Figure 2**).

**Figure 2.**
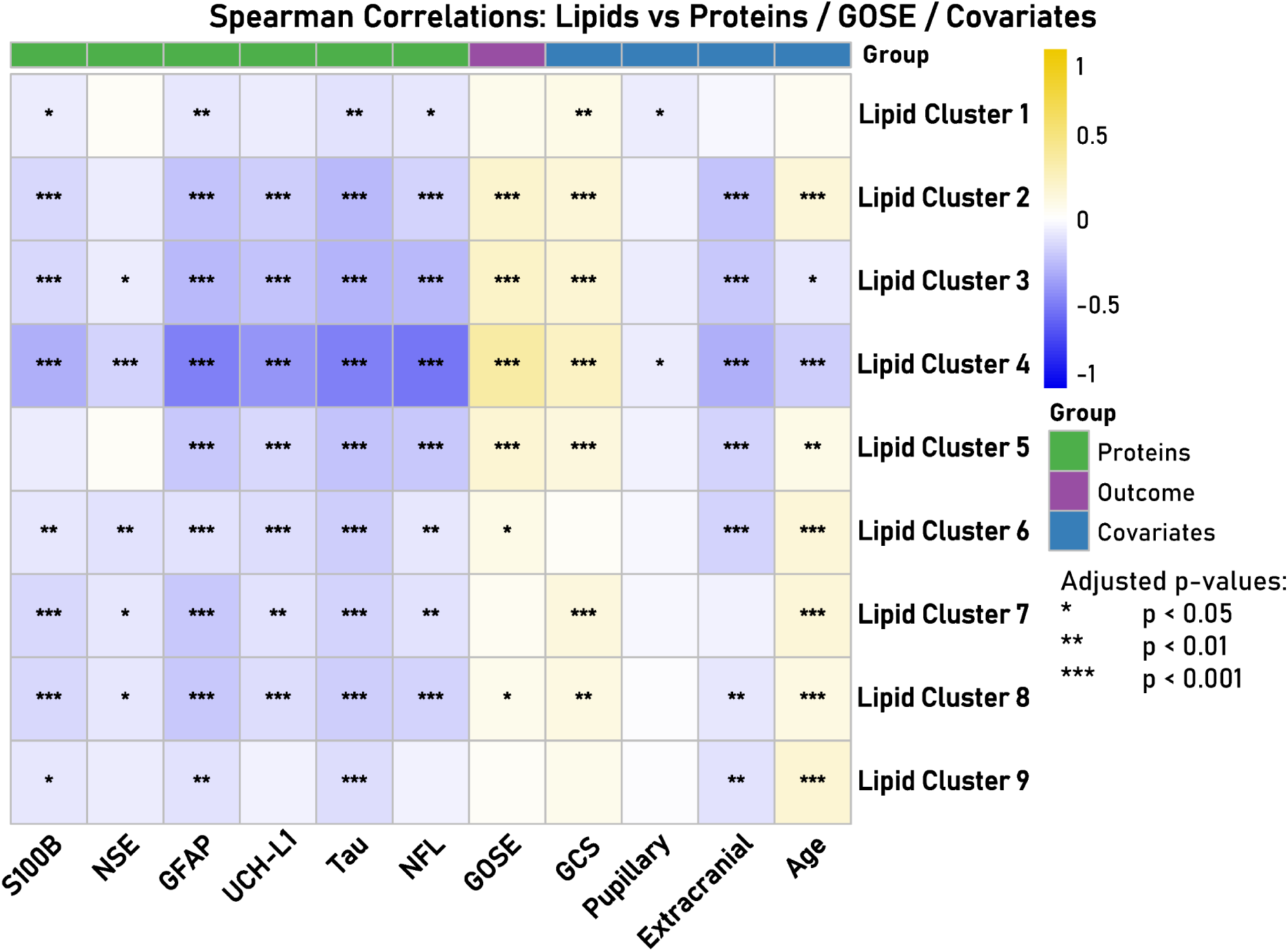
Correlation analysis between lipid clusters, protein biomarkers and clinical covariates. Colour scale displays Spearman’s rank correlation coefficient (ρ). p-value significances were adjusted using Benjamini-Hochberg false discovery rate (FDR). Significance levels: *** : p-value < 0.001; ** : p-value < 0.01; * : p-value < 0.05

The negative and positive correlations were markedly stronger for LC 4, which is comprised mostly of LPCs. LC 2 contains mostly SMs and Ceramides, while LC 3 contains mainly ether-linked PCs (PC-O). We also observed the same correlations between proteins and GOSE outcome scale to a lesser extent in LC 5, which contains mostly PCs in the cluster composition.

### Effect of choline in individual GOSE levels

The summed intensity of all choline-containing lipids increased monotonically across the GOSE scale in all 929 patients (Jonckheere–Terpstra p < 0.0001; **Figure 3**). The trend was evaluated on the full, uncollapsed 1–8 scale rather than on the five levels used for ordinal modelling and was visible across the whole range rather than being driven by the extremes. This association is unadjusted for injury severity.

**Figure 3.**
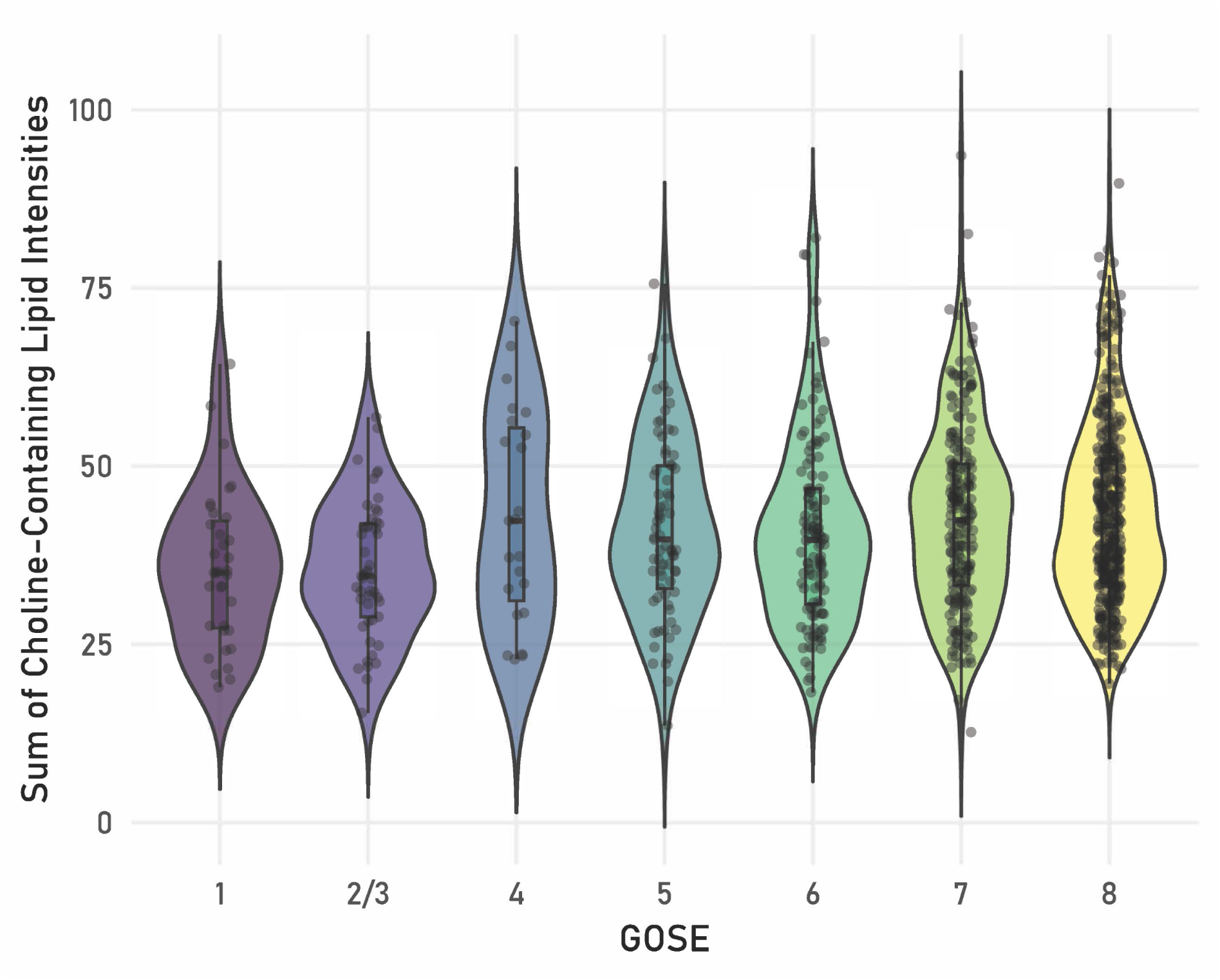
Sum of choline-containing lipids intensities for each GOSE level. Jonckheere-Terpstra trend test was applied to check possible trends over the outcome scale, which was statistically significant (p < 0.0001).

### Ordinal analysis shows incremental value of lipids in prediction models

In the model development set, regularised ordinal regression with CRASH ^25^ covariates as adjustment terms retained 10 variables in more than 70% of 20 stability runs: three proteins (S100B, GFAP, NfL), three annotated lipids (LPC(18:2), LPC(20:5), PC(O-34:3)) and four unannotated features (Unknown1, Unknown69, Unknown90, Unknown157). All ten were carried into the prediction model. The four unannotated features contribute to model performance but cannot be interpreted biologically and were therefore excluded from the trend analyses reported below.

Two proportional odds models were fitted in the development set: one with CRASH covariates alone, and one adding the 10 selected proteins and lipids. Both were then applied unchanged to the held-out test set. In the test set, the combined model gave a higher AUC than the CRASH-only model at every cumulative threshold (**Figure 4**), but the difference reached nominal significance at only one: GOSE ≥ 7 (AUC 0.728, 95% CI 0.671–0.785; DeLong p = 0.022). This p-value is unadjusted; after correction for the four threshold comparisons (Benjamini–Hochberg) no difference remained significant, and this result should therefore be read as exploratory. The AUC values for the baseline model containing CRASH variables only, the AUC for the baseline with proteins and lipids added, Delta AUC, unadjusted p-value and adjusted p-values are shown in **Table S2**. Averaged across thresholds, the dichotomous c-index in the test set rose from 0.720 (95% CI 0.672–0.767) for CRASH alone to 0.751 (0.702–0.798) with proteins and lipids added — an absolute gain of 0.031. Pseudo-Nagelkerke R^2^ rose from 0.125 for CRASH alone to 0.242 in the model with lipids and proteins added, yielding a gain of 0.117. When analysing the supplementary models (CRASH plus proteins-only and CRASH plus lipids-only, **Table S3**), while we observed a better discrimination for the proteins-only model, the lipids-only model presented a higher explained variance. Interestingly, none of these models alone was significant on their own in any threshold, which was true for the combined panel for GOSE ≥ 7.

**Figure 4.**
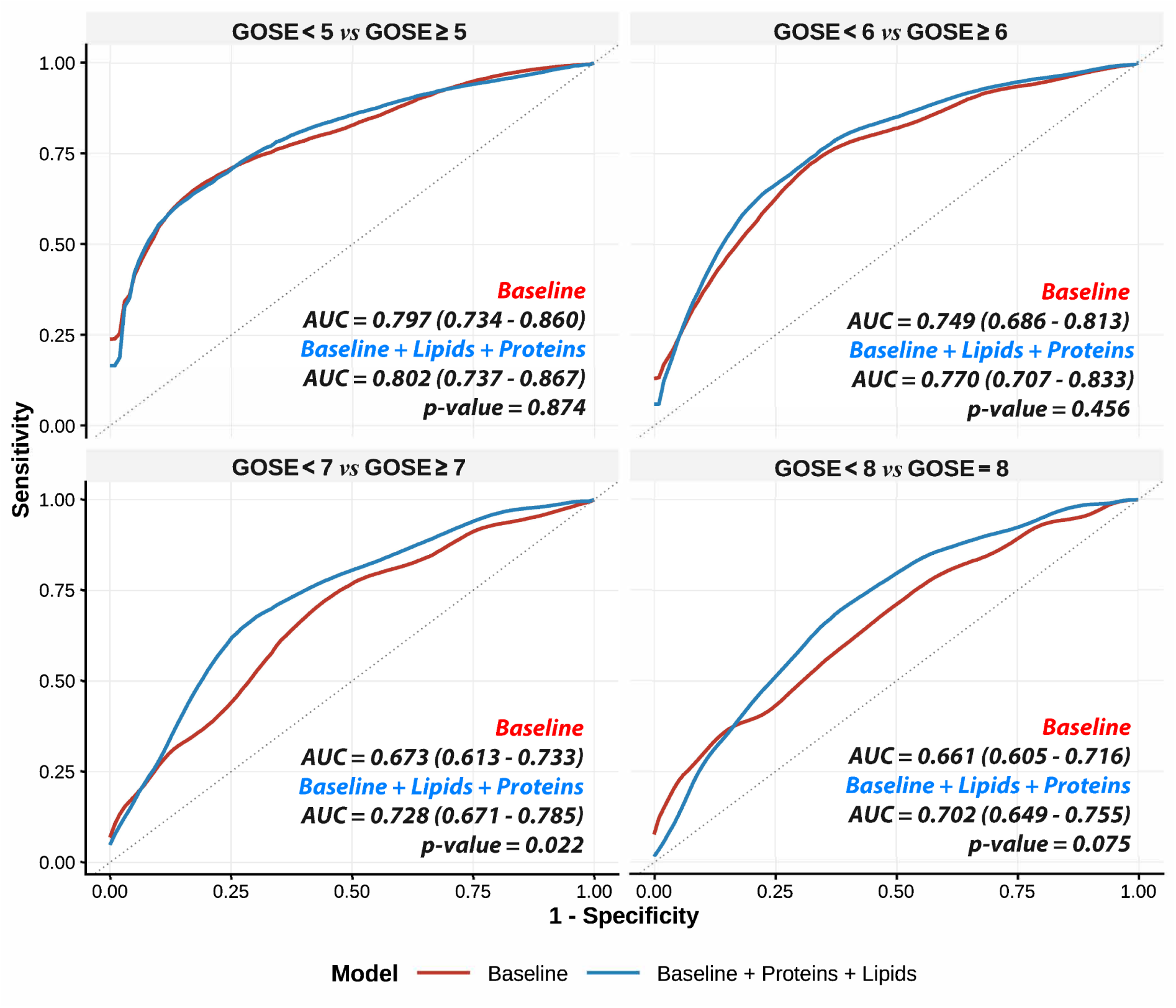
ROC Curves for the proportional odds logistic regression models. CRASH covariates were used as a baseline, CRASH covariates plus lipids and proteins were used as features for model building.

Results from the multiple imputation sensitivity analysis were consistent with the results from the complete case analysis, showing that using the complete cases instead of imputed covariates did not change the associations observed. The direction and magnitude for the selected variables were mostly preserved after imputation after checking the ORs from each variable (**Table S4**). ORs represent the change in odds of being in a higher GOSE category associated with a one unit increase in a chosen predictor, while keeping all other variables constant^33^.

Estimates for GCS and Pupillary reactiveness were directionally consistent or had overlapping confidence intervals, however, their confidence intervals were wider than those from the selected proteins and lipids. Overall, applying multiple imputation on the covariates that presented missing values did not majorly altered the principal associations performed in the complete case analysis.

### Trend test for ordinal-net selected and annotated lipids and proteins

The three annotated lipids and the three proteins were examined individually across the full GOSE scale in all 929 patients (**Figure 5**). The lipids increased with better outcome and the proteins decreased, all with Jonckheere–Terpstra p < 0.001. The opposing direction of the two marker types is the substantive observation here; these are unadjusted univariable trends and do not account for injury severity or for one another.

**Figure 5.**
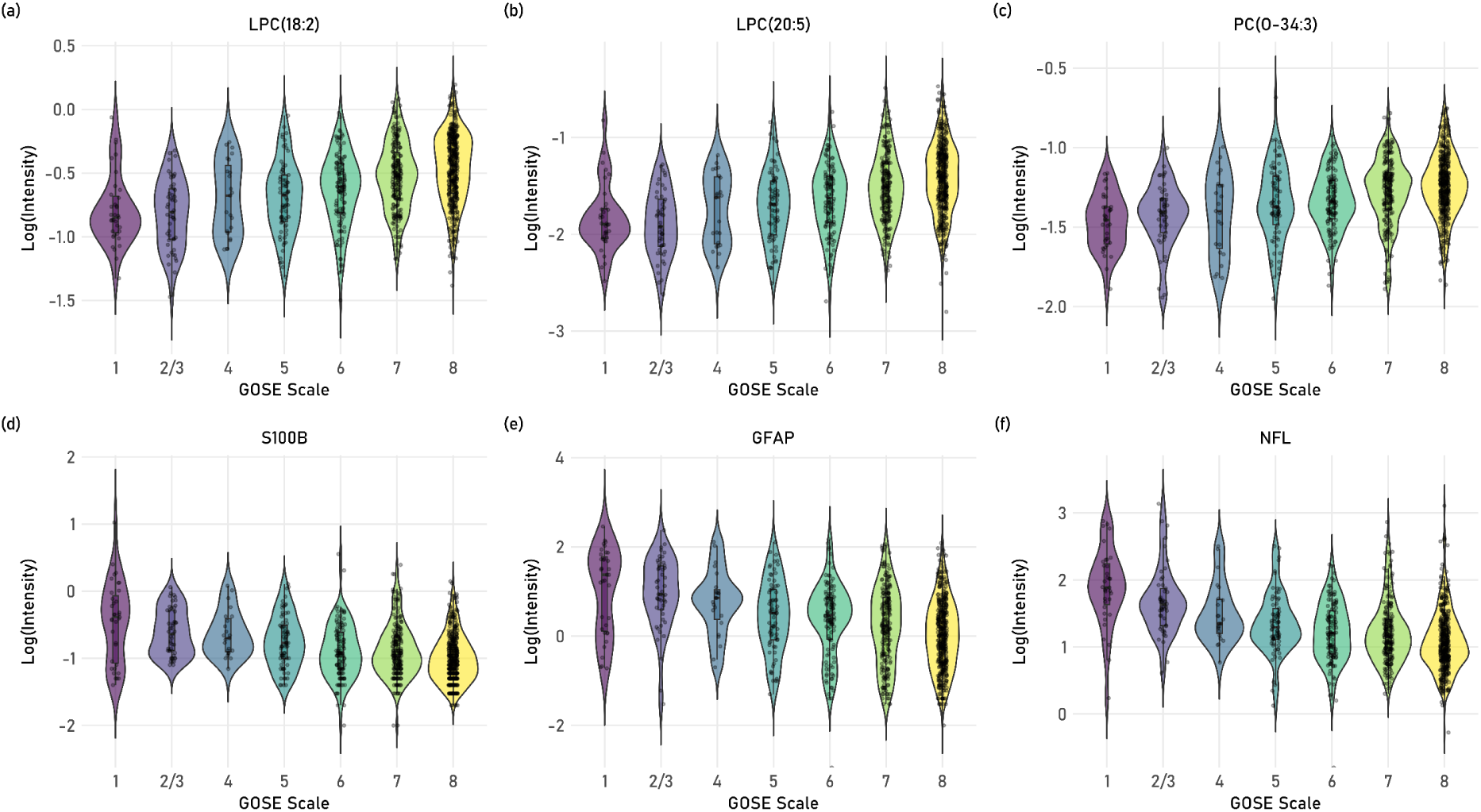
Ordinal Elastic-Net selected proteins and annotated lipid levels over the GOSE outcome scale. (LPC(18:2) **(a)**, LPC(20:5) **(b)**, PC(O-34:3) **(c),** S100B **(d)**, GFAP **(e)**, NfL **(f)**). All lipids and proteins presented a p-value < 0.001 in the Jonckheere-Terpstra trend test.

## Discussion

In this study, we investigated the serum lipids in patients from the CENTER-TBI cohort,^2^ more specifically patients with TBI that presented a GCS between 13-15. The circulating lipidome tracks 6-month functional outcome across the whole GOSE range; lipids and injury proteins move in opposite directions and are selected together rather than in place of one another; and the incremental prognostic value of the combined panel, though small, is concentrated at the top end of the GOSE ordinal scale, identifying patients who made a good recovery rather than those left with moderate disability or worse.

Although several studies have presented metabolomics-based prediction models for TBI in human samples, none has focused specifically on mTBI in this setting, instead either mixing mTBI with TBI cases presenting higher GCS score,^22,34–37^ focusing solely on more severe cases,^38^ or only analysing one single group setting.^39^ Our results indicate that circulating lipids carry outcome-related information across the GOSE scale, with a small but measurable incremental contribution that is concentrated at the upper recovery thresholds. To our knowledge, this is also the first study to model proteins and lipids jointly as predictors in mTBI, and to do so against the full ordinal outcome scale rather than a dichotomised endpoint.

Looking at the lipids contained in each of the most important Lipid Clusters in our clustering analysis, it is noteworthy the presence of SM, Ceramides, LPCs and ether PCs (PC-O), and their negative association with well-known protein biomarkers (GFAP, UCH-L1, T-TAU, S100B and NfL), together with a positive association with the GOSE outcome scale. Protein biomarkers are known for displaying higher levels in worse outcomes,^11–13,40^ which, depending on the individual marker, may reflect astrocytic damage, axonal degeneration and inflammatory processes.^41–44^ On the other hand, lipids were already reported as presenting higher levels in good outcomes,^19,22,39^ which can reflect the activation of damage repair pathways on the brain, such as the Lands pathway and CDP-Choline pathway.^45–48^

The direction of these associations raises a question that matters clinically: is the lipid signal simply a downstream marker of how severe the injury was, or does it carry information beyond injury severity? Patients who recover well will, on average, have sustained a lesser insult, and the lipid changes could be a proxy for that. Three observations argue that this is not the whole explanation. First, the lipid-outcome associations persisted after adjustment for the CRASH covariates, which capture admission GCS, pupillary reactivity, extracranial injury and age. Second, the selected lipids and the selected proteins moved in opposite directions across the GOSE scale, which argues against the lipids being simple markers of tissue injury and is more consistent with a marker that integrates host response, resilience and, potentially, recovery capacity. Third, elastic-net retained lipids alongside the proteins rather than in place of them, implying non-redundant information. None of this establishes causation. Our design is observational and single-timepoint, adjustment for CRASH is adjustment for a coarse severity summary rather than for severity itself, and residual confounding by injury burden is likely. The lipid signature should therefore be read as prognostically associated with recovery, not as a mechanism of it.

A closely related caveat is specificity. Low circulating LPC is not a brain-specific finding: LPC concentrations fall in sepsis and other critical illness and correlate inversely with mortality in those settings,^49^ and depressed LPC is a fairly general feature of systemic inflammation. Almost a quarter of our cohort had extracranial injuries, and low LPC in patients with poor outcome may in part reflect overall injury and inflammatory burden rather than a brain-specific deficit in membrane repair. Distinguishing these possibilities requires comparison against trauma controls without head injury, and against patients with systemic inflammation and no trauma - comparisons that CENTER-TBI, which recruited only patients with TBI, cannot supply. Until such data exist, the LPC and PC-O findings should be regarded as prognostically useful but mechanistically unresolved, and any future assay would need to be interpreted in the light of concurrent systemic illness.

Previously we observed correlations in choline-containing lipid with outcome severity, with increased values in mild TBI patients,^22^ and interestingly, we observed the same pattern in this work, which focused only in the mild cases. Using a Jonckheere-Terpstra test, we analysed if there was a significant trend in the sum of the intensities of all choline-containing lipids among the outcome scale, showing that this trend is statistically significant. Other studies point to an elevation of choline in brain after TBI,^50–52^ which may indicate cellular damage due to membrane disruption after TBI as a result of astrocytosis or inflammation,^52^ which at first may seem counterintuitive, however, the causes and locations of these dysregulations are different. Choline increase happens in brain and is due to membrane disruption and activation of phospholipases, which leads to the breakdown of choline-containing phospholipids, yielding choline and free fatty acids,^48,53^ while choline-containing phospholipid increase is detected in the circulating blood, where it reflects the transport of these lipids to the brain for membrane repair, since PC-containing lipids are synthesised in the liver and secreted as components of very low density lipoprotein (VLDL),^45^ then used as substrate to generate PCs and PEs for membrane repair.^46^

Given the nature of untargeted metabolomics and lipidomics datasets, containing hundreds (sometimes even thousands) of variables, it is important to select among them the variables that are the most important statistically for the biological question of the study. Also, given the ordinal scale of the GOSE outcome scale, it is important to use methods that are appropriate to deal with the ordinality of the present data. With that in mind, we chose to use a regularised ordinal regression model to select the variables that are mostly associated to the ordinal scale. Based on the ordinal logistic regression results, 3 of the 7 selected lipids were annotated (LPC(18:2), LPC(20:5), PC(O-34:3)), and the same lipids were selected in our earlier analysis of this cohort, which used a subset of the present development set and a different statistical approach.^22^ LPC (18:2) was also reported as increased together with other LPCs and LPEs in better outcomes in mTBI patients dichotomised as good (GOSE ≥ 7) or bad (GOSE ≤ 6) after 6 months of injury.^39^ LPC(20:5) is an interesting case to analyse individually due to the fatty acid chain that it carries, eicosapentaenoic acid (EPA), which has been investigated on its potential to lessen the secondary effects of TBI in brain and ameliorate inflammation.^53–55^ In another of our previous studies, we observed a strong correlation between LPC(18:2) and LPC(20:5) and brain abnormalities in magnetic resonance imaging, more specifically with fractional anisotropy, which is an imaging marker of white matter injury, indicating a possible relationship of LPCs and inflammatory response or myelin breakdown.^56–58^ PC(O-34:3) is an ether phosphatidylcholine, differing from a conventional PC by the presence of an ether bond rather than an ester bond. They constitute around 20% of the total phospholipid pool, and can be found in brain, heart, spleen and white blood cells, being involved in several biological functions such as cellular signalling, oxidative stress reduction and structural functions in the cell membrane.^59^ In the brain, these lipids perform important structural activities, and the deficiency of these lipids may be linked to neurodegeneration and defective myelin formation.^60^ It is known that TBI can cause white matter injury, and that the extent of this injury is determinant in the long-term effects of TBI. Axonal injury in the white matter can lead to demyelination, and the repair mechanisms that are responsible for rebuilding the myelin sheet in the axons might be impaired in patients with worse outcomes,^57^ thus reflecting the lower levels of these lipids in those patients.

Aside from the chosen lipids, the ordinal model also chose 3 protein biomarkers: GFAP, NfL and S100B. These proteins have been studied and reviewed extensively over the last years as potential biomarkers in TBI to predict functional outcomes, CT abnormalities, mortality and structural markers as a complement to other prognostic models based on clinical variables, especially since these models perform poorly in mTBI cases.^11,15,40,61^ GFAP and S100B are two astrocytic proteins that are released after TBI injury and may serve as severity markers,^41^ and possess different functions in brain. GFAP is a monomeric filament protein that has a cytoskeleton support function, while S100B regulates several different cell activities and cell morphology processes, including repair and tissue development, and the increase of these two proteins may be indicative of damage in the Blood-Brain barrier (BBB), which can happen even in subconcussive and non-concussive injuries.^62^ NfL on the other hand, is a structural component of the neuronal cytoplasm which gives stability to neurons and expressed mainly in long myelinated white matter axons.^3,12^ NfL levels in plasma after injury are acutely elevated after severe and moderate TBI,^63^ while in mTBI these levels might reach its maxima only at a later sampling time,^61^ and alterations in this protein levels may indicate traumatic axonal injury.^3^ One interesting aspect to consider when we observe the variables selected is the fact that it selected proteins and lipids together in the same model, picking together well known injury markers (proteins) and potential lipid markers that can be related to repair mechanisms after injury, therefore choosing a composite panel that covers both sides of TBI: injury and recovery.

We also evaluated how the selected proteins and lipids performed in predicting 6-month outcome in patients with a high GCS, benchmarking them against a panel of CRASH variables - a well-established set of clinical predictors validated for moderate and severe TBI. CRASH is known to underperform in mTBI, and our aim was to test whether adding proteins and lipids could improve prediction in this group. Combining CRASH variables with proteins and lipids increased the AUC at every threshold. However, the increment in AUC was not uniform through the GOSE ordinal range - it was least at the GOSE 4 / 5 interface, and maximal at the GOSE 6 / 7 and 7 / 8 interface. The improvement was modest in absolute terms, a gain of about 0.03 in the averaged c-index, and, once the four threshold comparisons are accounted for, no single threshold difference remains statistically significant. What can reasonably be claimed is narrower than statistical significance at one cut-point would suggest: the selected lipids and proteins contribute complementary information to clinical variables, and that contribution is largest where CRASH is weakest, namely at the upper end of the outcome scale, where good recovery is separated from moderate disability and below. This is a signal worth pursuing, not a model ready for use.

Taken together, circulating lipids are informative in mTBI and correlate with clinical variables and with established protein biomarkers. We further identified a clear trend in serum choline-containing lipids, with lower levels in patients with worse outcomes. Using ordinal elastic-net logistic regression, we selected a combined panel of proteins and lipids that captures both injury markers and candidate repair markers, verifying their prediction potential using proportional odds logistic regression. The main advantage of using the *polr* model is that this model uses the full ordinal scale, instead of dichotomising the outcomes in “good” or “bad”. An ordinal model, rather than a dichotomised one, provides greater statistical power for a given sample size,^64–66^ while also capturing meaningful differences across ordinal outcomes scales as a whole.^67^. Analysing the outcome as an ordinal scale, rather than as dichotomised endpoints, identifies variables that better explain the GOSE scale as a whole and avoids the bias introduced by arbitrary cut-points. This is relevant given ongoing debate about the GOSE scale itself, a summary measure with broad categories that does not discriminate well between mental and physical disability,^68^ and analysing the scale as a whole, rather than at dichotomous cut-points, allows a molecular signal to be related to the full range of recovery states. Whether that translates into better clinical decisions is a separate question, and one our data cannot answer: the gains we observed were small and confined to a single threshold.

The number of patients with poor outcome was small (GOSE ≤ 4, n = 107; 11.5%), as is expected in an mTBI cohort in which most patients cluster at the upper end of the scale (GOSE 7 and 8). This class imbalance constrains model fitting and required us to collapse all patients with GOSE ≤ 4 into a single level (GOSE 1-4). Samples were also collected within 24 h of injury, which may blunt the contribution of some protein biomarkers, which are reported as having a maximum at different time points after injury, such as S100B and NfL.^61,69^ Besides, we analysed together with the lipids only 6 protein biomarkers which were already quantified previously by the CENTER-TBI cohort. A comprehensive multi-omics study of lipidomics, metabolomics and proteomics, as well as cytokines would possibly be able to illustrate the full extent of potential injury and recovery markers in TBI, improving outcome prediction and TBI management. Also, the lipidomics analyses were restricted to positive ion mode only, excluding other potential lipids that could provide additional information.

Analyses were restricted to patients with complete data for the variables entering each model, and missing values were not imputed. This excluded 151 of the 929 patients (16.3%) from the modelling step; 92 in the development set and 59 in the test set. Because completeness is unlikely to be independent of injury severity and care pathway, this may have introduced selection bias. Even though the sensitivity analysis comparing the complete cases with the imputed data showed robustness to some extent in the complete case analysis, having the complete information at hand for all patients would be better to fully describe the clinical condition of the entire cohort and improve statistical power.

Comparisons of model discrimination were made at four cumulative GOSE thresholds without pre-specified adjustment for multiplicity. The single nominally significant result, at GOSE ≥ 7, does not survive Bonferroni, Holm–Bonferroni or Benjamini–Hochberg correction. We report it because it is the threshold we had reason to expect on clinical grounds and because unadjusted threshold-wise comparisons are the convention in this literature, but it must be treated as exploratory rather than confirmatory.

We also lacked a trauma comparison group without head injury, so we cannot separate a brain-specific lipid response from a general response to injury and inflammation. The lipids were measured on an untargeted, semi-quantitative research platform, and four of the seven selected lipid features remain structurally unannotated; neither is compatible with immediate clinical deployment. Finally, CENTER-TBI recruited hospital-presenting patients in Europe, with a mean age of 52 years and a high proportion of ground-level falls; these findings should not be extrapolated to sports-related, military or non-hospital-presenting mTBI without direct study.

Two aspects of these results are potentially actionable. First, the incremental information provided by the lipid markers was not uniform across the GOSE scale: the benefit of adding lipids was minimal at the lower end (the GOSE 4/5 interface) and greatest at the top (the GOSE 6/7 and 7/8 interfaces). It is the top of the scale that matters most in this population, because the clinical question at discharge is not survival or dependency but whether a patient who looks well will in fact return to their previous work, driving and family life. It is also the decision that existing tools handle worst, because CRASH excluded patients with a GCS of 15, and IMPACT was developed on patients with moderate or severe TBI. A marker that helps flag the patient at risk of incomplete recovery would address a real gap in a system where fewer than one in ten patients discharged after mTBI is seen again.

Second, the inverse relationship between the protein biomarkers and the metabolic markers suggests that injury severity and metabolic host response carry complementary information. The latter arguably integrates individual host resilience and may offer a different perspective on recovery potential. If that separation holds in independent data, it points to a different clinical use for blood-based testing in mTBI than the established one. GFAP and UCH-L1 are used on the day of injury to decide who does not need a CT scan;^17^ a repair-associated marker would instead help decide who needs to be seen again in the weeks that follow. These are complementary, not competing, applications.

Several concrete steps stand between these findings and any clinical use. The lipids reported here would need to be measured with targeted, quantitative assays with defined reference intervals and documented analytical performance, and the four unannotated features would need structural identification before a panel could be standardised. The findings require replication in a genuinely independent cohort and sampled at a defined interval after injury rather than within a 24-hour window. Because the absolute gain in discrimination was small, replication should be judged not by statistical significance but by clinical utility — through calibration and decision-curve analysis, and ultimately through a study in which follow-up is actually allocated on the basis of the marker and patient outcomes are compared. Serial sampling would also help separate the injury and repair components, which a single early timepoint cannot.

In 929 patients with mTBI from CENTER-TBI, the acute serum lipidome was associated with 6-month functional outcome across the full ordinal GOSE scale. Lysophosphatidylcholines and ether-linked phosphatidylcholines rose with better recovery while astroglial and axonal injury proteins fell, and the two marker classes were selected together, suggesting they capture complementary aspects of injury and repair. Adding them to clinical variables improved discrimination modestly, with the gain concentrated at the upper end of the outcome scale, where good recovery is separated from moderate disability and below. The effect is small and requires replication with quantitative assays before it can inform care, but it identifies a clinically meaningful target: recognising, at presentation, the patient with apparently mild injury who will not fully recover.

## Data availability

The lipidomics data are stored at the department of Advanced Data Management at Leiden University Medical Center (LUMC; Leiden, NL) and available for researchers upon submission of a data access request through the CENTER-TBI website: https://www.center-tbi.eu/data. The authors are not legally allowed to share it publicly. The authors confirm that they received no special access privileges to the data.

CENTER-TBI is committed to data sharing, and in particular to responsible further use of the data. Hereto, we have a data sharing statement in place: https://www.center-tbi.eu/data/sharing. The CENTER-TBI Management Committee, in collaboration with the General Assembly, established the Data Sharing policy and Publication and Authorship Guidelines to assure correct and appropriate use of the data as the dataset is hugely complex and requires help of experts from the Data Curation Team or Bio-Statistical Team for correct use. This means that we encourage researchers to contact the CENTER-TBI team for any research plans and the Data Curation Team for any help in appropriate use of the data, including sharing of scripts. The complete Manual for data access is also available online: https://www.center-tbi.eu/files/SOP-Manual-DAPR-20181101.pdf

## Data Availability

The lipidomics data are stored at the department of Advanced Data Management at Leiden University Medical Center (LUMC; Leiden, NL) and available for researchers upon submission of a data access request through the CENTER-TBI website: https://www.center-tbi.eu/data. The authors are not legally allowed to share it publicly. The authors confirm that they received no special access privileges to the data.
CENTER-TBI is committed to data sharing, and in particular to responsible further use of the data. Hereto, we have a data sharing statement in place: https://www.center-tbi.eu/data/sharing. The CENTER-TBI Management Committee, in collaboration with the General Assembly, established the Data Sharing policy and Publication and Authorship Guidelines to assure correct and appropriate use of the data as the dataset is hugely complex and requires help of experts from the Data Curation Team or Bio-Statistical Team for correct use. This means that we encourage researchers to contact the CENTER-TBI team for any research plans and the Data Curation Team for any help in appropriate use of the data, including sharing of scripts. The complete Manual for data access is also available online: https://www.center-tbi.eu/files/SOP-Manual-DAPR-20181101.pdf

## Acknowledgments

The authors would like to thank Ewout W. Steyerberg, Youngjune Bhak and Romit Samanta for providing data and statistical support.

## Appendix 1

### The CENTER-TBI participants and investigators

Cecilia Åkerlund^1^, Krisztina Amrein^2^, Nada Andelic^3^, Lasse Andreassen^4^, Audny Anke^5^, Anna Antoni^6^, Gérard Audibert^7^, Philippe Azouvi^8^, Maria Luisa Azzolini^9^, Ronald Bartels^10^, Pál Barzó^11^, Romuald Beauvais^12^, Ronny Beer^13^, Bo-Michael Bellander^14^, Antonio Belli^15^, Habib Benali^16^, Maurizio Berardino^17^, Luigi Beretta^9^, Morten Blaabjerg^18^, Peter Bragge^19^, Alexandra Brazinova^20^, Vibeke Brinck^21^, Joanne Brooker^22^, Camilla Brorsson^23^, Andras Buki^24^, Monika Bullinger^25^, Manuel Cabeleira^26^, Alessio Caccioppola^27^, Emiliana Calappi ^27^, Maria Rosa Calvi^9^, Peter Cameron^28^, Guillermo Carbayo Lozano^29^, Marco Carbonara^27^, Simona Cavallo^17^, Giorgio Chevallard^30^, Arturo Chieregato^30^, Giuseppe Citerio^31, 32^, Hans Clusmann^33^, Mark Coburn^34^, Jonathan Coles^35^, Jamie D. Cooper^36^, Marta Correia^37^, Amra Čović ^38^, Nicola Curry^39^, Endre Czeiter^40^, Marek Czosnyka^26^, Claire Dahyot-Fizelier^41^, Paul Dark^42^, Helen Dawes^43^, Véronique De Keyser^44^, Vincent Degos^16^, Francesco Della Corte^45^, Hugo den Boogert^10^, Bart Depreitere^46^, Đula Đilvesi ^47^, Abhishek Dixit^48^, Emma Donoghue^22^, Jens Dreier^49^, Guy-Loup Dulière^50^, Ari Ercole^48^, Patrick Esser^43^, Erzsébet Ezer^51^, Martin Fabricius^52^, Valery L. Feigin^53^, Kelly Foks^54^, Shirin Frisvold^55^, Alex Furmanov^56^, Pablo Gagliardo^57^, Damien Galanaud^16^, Dashiell Gantner^28^, Guoyi Gao^58^, Pradeep George^59^, Alexandre Ghuysen^60^, Lelde Giga^61^, Ben Glocker^62^, Jagoš Golubovic^47^, Pedro A. Gomez ^63^, Johannes Gratz^64^, Benjamin Gravesteijn^65^, Francesca Grossi^45^, Russell L. Gruen^66^, Deepak Gupta^67^, Juanita A. Haagsma^65^, Iain Haitsma^68^, Raimund Helbok^69,70^, Eirik Helseth^71^, Lindsay Horton ^72^, Jilske Huijben^65^, Peter J. Hutchinson^73^, Bram Jacobs^74^, Stefan Jankowski^75^, Mike Jarrett^21^, Ji-yao Jiang^59^, Faye Johnson^76^, Kelly Jones^53^, Mladen Karan^47^, Angelos G. Kolias^73^, Erwin Kompanje^77^, Daniel Kondziella^52^, Evgenios Kornaropoulos^48^, Lars-Owe Koskinen^78^, Noémi Kovács^79^, Ana Kowark^80^, Alfonso Lagares^63^, Linda Lanyon^59^, Steven Laureys^81^, Fiona Lecky^82, 83^, Didier Ledoux^81^, Rolf Lefering^84^, Valerie Legrand^85^, Aurelie Lejeune^86^, Leon Levi^87^, Roger Lightfoot^88^, Hester Lingsma^65^, Andrew I.R. Maas^44,89^, Ana M. Castaño-León^63^, Marc Maegele^90^, Marek Majdan^20^, Alex Manara^91^, Geoffrey Manley^92^, Costanza Martino^93^, Hugues Maréchal^50^, Julia Mattern^94^, Catherine McMahon^95^, Béla Melegh^96^, David Menon^48^, Tomas Menovsky^44,89^, Ana Mikolic^65^, Benoit Misset^81^, Visakh Muraleedharan^59^, Lynnette Murray^28^, Ancuta Negru^97^, David Nelson^1^, Virginia Newcombe^48^, Daan Nieboer^65^, József Nyirádi^2^, Otesile Olubukola^82^, Matej Oresic^98^, Fabrizio Ortolano^27^, Aarno Palotie^99, 100, 101^, Paul M. Parizel^102^, Jean-François Payen^103^, Natascha Perera^12^, Vincent Perlbarg^16^, Paolo Persona^104^, Wilco Peul^105^, Anna Piippo-Karjalainen^106^, Matti Pirinen^99^, Dana Pisica^65^, Horia Ples^97^, Suzanne Polinder^65^, Inigo Pomposo^29^, Jussi P. Posti ^107^, Louis Puybasset^108^, Andreea Radoi ^109^, Arminas Ragauskas^110^, Rahul Raj^106^, Malinka Rambadagalla^111^, Isabel Retel Helmrich^65^, Jonathan Rhodes^112^, Sylvia Richardson^113^, Sophie Richter^48^, Samuli Ripatti^99^, Saulius Rocka^110^, Cecilie Roe^114^, Olav Roise^115,116^, Jonathan Rosand^117^, Jeffrey V. Rosenfeld^118^, Christina Rosenlund^119^, Guy Rosenthal^56^, Rolf Rossaint^80^, Sandra Rossi^104^, Daniel Rueckert^62^ Martin Rusnák^120^, Juan Sahuquillo^109^, Oliver Sakowitz^94, 121^, Renan Sanchez-Porras^121^, Janos Sandor^122^, Nadine Schäfer^84^, Silke Schmidt^123^, Herbert Schoechl^124^, Guus Schoonman^125^, Rico Frederik Schou^126^, Elisabeth Schwendenwein^6^, Charlie Sewalt^65^, Ranjit D. Singh^105^, Toril Skandsen^127, 128^, Peter Smielewski^26^, Abayomi Sorinola^129^, Emmanuel Stamatakis^48^, Simon Stanworth^39^, Robert Stevens^130^, William Stewart^131^, Ewout W. Steyerberg^65, 132, 133^, Nino Stocchetti^134^, Nina Sundström^135^, Riikka Takala^136^, Viktória Tamás^129^, Tomas Tamosuitis^137^, Mark Steven Taylor^20^, Aurore Thibaut^81^, Braden Te Ao^53^, Olli Tenovuo^107^, Alice Theadom^53^, Matt Thomas^91^, Dick Tibboel^138^, Marjolein Timmers^77^, Christos Tolias^139^, Tony Trapani^28^, Cristina Maria Tudora^97^, Andreas Unterberg^94^, Peter Vajkoczy ^140^, Shirley Vallance^28^, Egils Valeinis^61^, Zoltán Vámos^51^, Mathieu van der Jagt^141^, Gregory Van der Steen^44^, Joukje van der Naalt^74^, Jeroen T.J.M. van Dijck^105^, Inge A. M. van Erp^105^, Thomas A. van Essen^105^, Wim Van Hecke^142^, Caroline van Heugten^143^, Ernest van Veen^65^, Thijs Vande Vyvere^144^, Roel P. J. van Wijk^105^, Alessia Vargiolu^32^, Emmanuel Vega^86^, Kimberley Velt^65^, Jan Verheyden^142^, Paul M. Vespa^145^, Anne Vik^127, 146^, Rimantas Vilcinis^137^, Victor Volovici^68^, Nicole von Steinbüchel^38^, Daphne Voormolen^65^, Petar Vulekovic^47^, Kevin K.W. Wang^147^, Daniel Whitehouse^48^, Eveline Wiegers^65^, Guy Williams^48^, Lindsay Wilson^72^, Stefan Winzeck^48^, Stefan Wolf^148^, Zhihui Yang^117^, Peter Ylén^149^, Alexander Younsi^94^, Marina Zeldovich^150^, Frederick A. Zeiler^48,151^, Veronika Zelinkova^20^, Agate Ziverte^61^, Tommaso Zoerle^27^

^1^ Department of Physiology and Pharmacology, Section of Perioperative Medicine and Intensive Care, Karolinska Institutet, Stockholm, Sweden

^2^ János Szentágothai Research Centre, University of Pécs, Pécs, Hungary

^3^ Division of Clinical Neuroscience, Department of Physical Medicine and Rehabilitation, Oslo University Hospital and University of Oslo, Oslo, Norway

^4^ Department of Neurosurgery, University Hospital Northern Norway, Tromso, Norway

^5^ Department of Physical Medicine and Rehabilitation, University Hospital Northern Norway, Tromso, Norway

^6^ Trauma Surgery, Medical University Vienna, Vienna, Austria

^7^ Department of Anesthesiology & Intensive Care, University Hospital Nancy, Nancy, France

^8^ Raymond Poincare hospital, Assistance Publique – Hopitaux de Paris, Paris, France

^9^ Department of Anesthesiology & Intensive Care, S Raffaele University Hospital, Milan, Italy

^10^ Department of Neurosurgery, Radboud University Medical Center, Nijmegen, The Netherlands

^11^ Department of Neurosurgery, University of Szeged, Szeged, Hungary

^12^ International Projects Management, ARTTIC, Munchen, Germany

^13^ Department of Neurology, Neurological Intensive Care Unit, Medical University of Innsbruck, Innsbruck, Austria

^14^ Department of Neurosurgery & Anesthesia & intensive care medicine, Karolinska University Hospital, Stockholm, Sweden

^15^ NIHR Surgical Reconstruction and Microbiology Research Centre, Birmingham, UK

^16^ Anesthesie-Réanimation, Assistance Publique – Hopitaux de Paris, Paris, France

^17^ Department of Anesthesia & ICU, AOU Città della Salute e della Scienza di Torino - Orthopedic and Trauma Center, Torino, Italy

^18^ Department of Neurology, Odense University Hospital, Odense, Denmark

^19^ BehaviourWorks Australia, Monash Sustainability Institute, Monash University, Victoria, Australia

^20^ Department of Public Health, Faculty of Health Sciences and Social Work, Trnava University, Trnava, Slovakia

^21^ Quesgen Systems Inc., Burlingame, California, USA

^22^ Australian & New Zealand Intensive Care Research Centre, Department of Epidemiology and Preventive Medicine, School of Public Health and Preventive Medicine, Monash University, Melbourne, Australia

^23^ Department of Surgery and Perioperative Science, Umeå University, Umeå, Sweden

^24^ Department of Neurosurgery, Örebro University and University Hospital, Örebro, Sweden

^25^ Department of Medical Psychology, Universitätsklinikum Hamburg-Eppendorf, Hamburg, Germany

^26^ Brain Physics Lab, Division of Neurosurgery, Dept of Clinical Neurosciences, University of Cambridge, Addenbrooke’s Hospital, Cambridge, UK

^27^ Neuro ICU, Fondazione IRCCS Cà Granda Ospedale Maggiore Policlinico, Milan, Italy

^28^ ANZIC Research Centre, Monash University, Department of Epidemiology and Preventive Medicine, Melbourne, Victoria, Australia

^29^ Department of Neurosurgery, Hospital of Cruces, Bilbao, Spain

^30^ NeuroIntensive Care, Niguarda Hospital, Milan, Italy

^31^ School of Medicine and Surgery, Università Milano Bicocca, Milano, Italy

^32^ NeuroIntensive Care Unit, Department Neuroscience, IRCCS Fondazione San Gerardo dei Tintori, Monza, Italy

^33^Department of Neurosurgery, Medical Faculty RWTH Aachen University, Aachen, Germany

^34^ Department of Anesthesiology and Intensive Care Medicine, University Hospital Bonn, Bonn, Germany

^35^ Department of Anesthesia & Neurointensive Care, Cambridge University Hospital NHS Foundation Trust, Cambridge, UK

^36^ School of Public Health & PM, Monash University and The Alfred Hospital, Melbourne, Victoria, Australia

^37^ Radiology/MRI department, MRC Cognition and Brain Sciences Unit, Cambridge, UK

^38^ Institute of Medical Psychology and Medical Sociology, Universitätsmedizin Göttingen, Göttingen, Germany

^39^ Oxford University Hospitals NHS Trust, Oxford, UK

^40^ Department of Neurosurgery, Medical School, University of Pécs, Hungary and Neurotrauma Research Group, János Szentágothai Research Centre, University of Pécs, Hungary

^41^ Intensive Care Unit, CHU Poitiers, Potiers, France

^42^ University of Manchester NIHR Biomedical Research Centre, Critical Care Directorate, Salford Royal Hospital NHS Foundation Trust, Salford, UK

^43^ Movement Science Group, Faculty of Health and Life Sciences, Oxford Brookes University, Oxford, UK

^44^ Department of Neurosurgery, Antwerp University Hospital, Edegem, Belgium

^45^ Department of Anesthesia & Intensive Care, Maggiore Della Carità Hospital, Novara, Italy

^46^ Department of Neurosurgery, University Hospitals Leuven, Leuven, Belgium

^47^ Department of Neurosurgery, Clinical centre of Vojvodina, Faculty of Medicine, University of Novi Sad, Novi Sad, Serbia

^48^ Division of Anaesthesia, University of Cambridge, Addenbrooke’s Hospital, Cambridge, UK

^49^ Center for Stroke Research Berlin, Charité – Universitätsmedizin Berlin, corporate member of Freie Universität Berlin, Humboldt-Universität zu Berlin, and Berlin Institute of Health, Berlin, Germany

^50^ Intensive Care Unit, CHR Citadelle, Liège, Belgium

^51^ Department of Anaesthesiology and Intensive Therapy, University of Pécs, Pécs, Hungary

^52^ Departments of Neurology, Clinical Neurophysiology and Neuroanesthesiology, Region Hovedstaden Rigshospitalet, Copenhagen, Denmark

^53^ National Institute for Stroke and Applied Neurosciences, Faculty of Health and Environmental Studies, Auckland University of Technology, Auckland, New Zealand

^54^ Department of Neurology, Erasmus MC, Rotterdam, the Netherlands

^55^ Department of Anesthesiology and Intensive care, University Hospital Northern Norway, Tromso, Norway

^56^ Department of Neurosurgery, Hadassah-hebrew University Medical center, Jerusalem, Israel

^57^ Fundación Instituto Valenciano de Neurorrehabilitación (FIVAN), Valencia, Spain

^58^ Department of Neurosurgery, Shanghai Renji hospital, Shanghai Jiaotong University/school of medicine, Shanghai, China

^59^ Karolinska Institutet, INCF International Neuroinformatics Coordinating Facility, Stockholm, Sweden

^60^ Emergency Department, CHU, Liège, Belgium

^61^ Neurosurgery clinic, Pauls Stradins Clinical University Hospital, Riga, Latvia

^62^ Department of Computing, Imperial College London, London, UK

^63^ Department of Neurosurgery, Hospital Universitario 12 de Octubre, Madrid, Spain

^64^ Department of Anesthesia, Critical Care and Pain Medicine, Medical University of Vienna, Austria

^65^ Department of Public Health, Erasmus Medical Center-University Medical Center, Rotterdam, The Netherlands

^66^ College of Health and Medicine, Australian National University, Canberra, Australia

^67^ Department of Neurosurgery, Neurosciences Centre & JPN Apex trauma centre, All India Institute of Medical Sciences, New Delhi-110029, India

^68^ Department of Neurosurgery, Erasmus MC, Rotterdam, the Netherlands

^69^ Department of Neurology, Kepler University Hospital, Johannes Kepler University Linz, Linz, Austria.

^70^ Clinical Research Institute for Neuroscience, Johannes Kepler University Linz, Linz, Austria

^71^ Department of Neurosurgery, Oslo University Hospital, Oslo, Norway

^72^ Division of Psychology, University of Stirling, Stirling, UK

^73^ Division of Neurosurgery, Department of Clinical Neurosciences, Addenbrooke’s Hospital & University of Cambridge, Cambridge, UK

^74^ Department of Neurology, University of Groningen, University Medical Center Groningen, Groningen, Netherlands

^75^ Neurointensive Care, Sheffield Teaching Hospitals NHS Foundation Trust, Sheffield, UK

^76^ Salford Royal Hospital NHS Foundation Trust Acute Research Delivery Team, Salford, UK

^77^ Department of Intensive Care and Department of Ethics and Philosophy of Medicine, Erasmus Medical Center, Rotterdam, The Netherlands

^78^ Department of Clinical Neuroscience, Neurosurgery, Umeå University, Umeå, Sweden

^79^ Hungarian Brain Research Program - Grant No. KTIA_13_NAP-A-II/8, University of Pécs, Pécs, Hungary

^80^ Department of Anaesthesiology, University Hospital of Aachen, Aachen, Germany

^81^ Cyclotron Research Center, University of Liège, Liège, Belgium

^82^ Centre for Urgent and Emergency Care Research (CURE), Health Services Research Section, School of Health and Related Research (ScHARR), University of Sheffield, Sheffield, UK

^83^ Emergency Department, Salford Royal Hospital, Salford UK

^84^ Institute of Research in Operative Medicine (IFOM), Witten/Herdecke University, Cologne, Germany

^85^ VP Global Project Management CNS, ICON, Paris, France

^86^ Department of Anesthesiology-Intensive Care, Lille University Hospital, Lille, France

^87^ Department of Neurosurgery, Rambam Medical Center, Haifa, Israel

^88^ Department of Anesthesiology & Intensive Care, University Hospitals Southhampton NHS Trust, Southhampton, UK

^89^ Department of Translational Neuroscience, Faculty of Medicine and Health Science, University of Antwerp, Antwerp, Belgium

^90^ Cologne-Merheim Medical Center (CMMC), Department of Traumatology, Orthopedic Surgery and Sportmedicine, Witten/Herdecke University, Cologne, Germany

^91^ Intensive Care Unit, Southmead Hospital, Bristol, Bristol, UK

^92^ Department of Neurological Surgery, University of California, San Francisco, California, USA

^93^ Department of Anesthesia & Intensive Care,M. Bufalini Hospital, Cesena, Italy

^94^ Department of Neurosurgery, University Hospital Heidelberg, Heidelberg, Germany

^95^ Department of Neurosurgery, The Walton centre NHS Foundation Trust, Liverpool, UK

^96^ Department of Medical Genetics, University of Pécs, Pécs, Hungary

^97^ Department of Neurosurgery, Emergency County Hospital Timisoara, Timisoara, Romania

^98^ School of Medical Sciences, Örebro University, Örebro, Sweden

^99^ Institute for Molecular Medicine Finland, University of Helsinki, Helsinki, Finland

^100^ Analytic and Translational Genetics Unit, Department of Medicine; Psychiatric & Neurodevelopmental Genetics Unit, Department of Psychiatry; Department of Neurology, Massachusetts General Hospital, Boston, MA, USA

^101^ Program in Medical and Population Genetics; The Stanley Center for Psychiatric Research, The Broad Institute of MIT and Harvard, Cambridge, MA, USA

^102^ Department of Radiology, University of Antwerp, Edegem, Belgium

^103^ Department of Anesthesiology & Intensive Care, University Hospital of Grenoble, Grenoble, France

^104^ Department of Anesthesia & Intensive Care, Azienda Ospedaliera Università di Padova, Padova, Italy

^105^ Dept. of Neurosurgery, Leiden University Medical Center, Leiden, The Netherlands and Dept. of Neurosurgery, Medical Center Haaglanden, The Hague, The Netherlands

^106^ Department of Neurosurgery, Helsinki University Central Hospital

^107^ Division of Clinical Neurosciences, Department of Neurosurgery and Turku Brain Injury Centre, Turku University Hospital and University of Turku, Turku, Finland

^108^ Department of Anesthesiology and Critical Care, Pitié -Salpêtrière Teaching Hospital, Assistance Publique, Hôpitaux de Paris and University Pierre et Marie Curie, Paris, France

^109^ Neurotraumatology and Neurosurgery Research Unit (UNINN), Vall d’Hebron Research Institute, Barcelona, Spain

^110^ Department of Neurosurgery, Kaunas University of technology and Vilnius University, Vilnius, Lithuania

^111^ Department of Neurosurgery, Rezekne Hospital, Latvia

^112^ Department of Anaesthesia, Critical Care & Pain Medicine NHS Lothian & University of Edinburg, Edinburgh, UK

^113^ Director, MRC Biostatistics Unit, Cambridge Institute of Public Health, Cambridge, UK

^114^ Department of Physical Medicine and Rehabilitation, Oslo University Hospital/University of Oslo, Oslo, Norway

^115^ Division of Orthopedics, Oslo University Hospital, Oslo, Norway

^116^ Institue of Clinical Medicine, Faculty of Medicine, University of Oslo, Oslo, Norway

^117^ Broad Institute, Cambridge MA Harvard Medical School, Boston MA, Massachusetts General Hospital, Boston MA, USA

^118^ National Trauma Research Institute, The Alfred Hospital, Monash University, Melbourne, Victoria, Australia

^119^ Department of Neurosurgery, Odense University Hospital, Odense, Denmark

^120^ International Neurotrauma Research Organisation, Vienna, Austria

^121^ Klinik für Neurochirurgie, Klinikum Ludwigsburg, Ludwigsburg, Germany

^122^ Division of Biostatistics and Epidemiology, Department of Preventive Medicine, University of Debrecen, Debrecen, Hungary

^123^ Department Health and Prevention, University Greifswald, Greifswald, Germany

^124^ Department of Anaesthesiology and Intensive Care, AUVA Trauma Hospital, Salzburg, Austria

^125^ Department of Neurology, Elisabeth-TweeSteden Ziekenhuis, Tilburg, the Netherlands

^126^ Department of Neuroanesthesia and Neurointensive Care, Odense University Hospital, Odense, Denmark

^127^ Department of Neuromedicine and Movement Science, Norwegian University of Science and Technology, NTNU, Trondheim, Norway

^128^ Department of Physical Medicine and Rehabilitation, St.Olavs Hospital, Trondheim University Hospital, Trondheim, Norway

^129^ Department of Neurosurgery, University of Pécs, Pécs, Hungary

^130^ Division of Neuroscience Critical Care, John Hopkins University School of Medicine, Baltimore, USA

^131^ Department of Neuropathology, Queen Elizabeth University Hospital and University of Glasgow, Glasgow, UK

^132^ Dept. of Department of Biomedical Data Sciences, Leiden University Medical Center, Leiden, The Netherlands

^133^ Julius Center for Health Sciences and Primary Care, University Medical Center Utrecht, Utrecht, The Netherlands

^134^ Department of Pathophysiology and Transplantation, Milan University, and Neuroscience ICU, Fondazione IRCCS Cà Granda Ospedale Maggiore Policlinico, Milano, Italy

^135^ Department of Radiation Sciences, Biomedical Engineering, Umeå University, Umeå, Sweden

^136^ Perioperative Services, Intensive Care Medicine and Pain Management, Turku University Hospital and University of Turku, Turku, Finland

^137^ Department of Neurosurgery, Kaunas University of Health Sciences, Kaunas, Lithuania

^138^ Intensive Care and Department of Pediatric Surgery, Erasmus Medical Center, Sophia Children’s Hospital, Rotterdam, The Netherlands

^139^ Department of Neurosurgery, Kings college London, London, UK

^140^ Neurologie, Neurochirurgie und Psychiatrie, Charité – Universitätsmedizin Berlin, Berlin, Germany

^141^ Department of Intensive Care Adults, Erasmus MC– University Medical Center Rotterdam, Rotterdam, the Netherlands

^142^ icoMetrix NV, Leuven, Belgium

^143^ Movement Science Group, Faculty of Health and Life Sciences, Oxford Brookes University, Oxford, UK

^144^ Radiology Department, Antwerp University Hospital and University of Antwerp, Edegem, Belgium

^145^ Director of Neurocritical Care, University of California, Los Angeles, USA

^146^ Department of Neurosurgery, St.Olavs Hospital, Trondheim University Hospital, Trondheim, Norway

^147^ Department of Emergency Medicine, University of Florida, Gainesville, Florida, USA

^148^ Department of Neurosurgery, Charité – Universitätsmedizin Berlin, corporate member of Freie Universität Berlin, Humboldt-Universität zu Berlin, and Berlin Institute of Health, Berlin, Germany

^149^ VTT Technical Research Centre, Tampere, Finland

^150^ Sigmund Freud University, Faculty of Psychotherapy Science, Vienna, Austria

^151^ Section of Neurosurgery, Department of Surgery, Rady Faculty of Health Sciences, University of Manitoba, Winnipeg, MB, Canada

