## Supplementary Material for "Serum lipidomics identifies outcome signatures in patients with traumatic brain injury, presenting with Glasgow Coma Scale score of 13-15"

### Informed consent and ethical approvals

The CENTER-TBI study (European Commission grant no. 602150) was conducted in agreement with all relevant laws of the European Union, and with local laws and regulations at the respective locations of 65 recruitment centres. A detailed description of the CENTER-TBI administrative, regulatory, and logistic framework is published elsewhere.[1] That publication also provides information regarding the data storage, de-identification, verification, and curation.

The CENTER-TBI study complied with relevant laws and regulations on the use of human materials, and all relevant guidance relating to clinical studies from time to time in force including, but not limited to, the ICH Harmonised Tripartite Guideline for Good Clinical Practice (CPMP/ICH/135/95) ("ICH GCP") and the World Medical Association Declaration of Helsinki entitled "Ethical Principles for Medical Research Involving Human Subjects".

Informed Consent by the patients and/or the legal representative/next of kin was obtained, accordingly to the local legislations, for all patients recruited in the Core Dataset of CENTER-TBI and documented in the e-CRF.

Ethical approval was obtained for each recruiting site. The list of sites, Ethical Committees, approval numbers and approval dates can be found on the website: https://www.center-tbi.eu/project/ethical-approval.

### Lipidomics analysis

10 µL of serum was extracted with 10 µL 0.9% NaCl and with 120 µL of $CHCl_3$: MeOH (2:1, v/v) solvent mixture containing internal standard mixture (c = 2.5 mg/mL; 1,2-diheptadecanoyl-sn-glycero-3-phosphoethanolamine (PE(17:0/17:0)), N-heptadecanoyl-D-erythro-sphingosylphosphorylcholine (SM(d18:1/17:0)), N-heptadecanoyl-D-erythro-sphingosine (Cer(d18:1/17:0)), 1,2-diheptadecanoyl-sn-glycero-3-phosphocholine (PC(17:0/17:0)), 1-heptadecanoyl-2-hydroxy-sn-glycero-3-phosphocholine (LPC(17:0)) and 1-palmitoyl-d31-2-oleoyl-sn-glycero-3-phosphocholine (PC(16:0/d31/18:1)) and, triheptadecanoylglycerol (TG(17:0/17:0/17:0)). The samples were vortexed and kept on ice for 30 min before centrifugation at

9400 rcf for 3 min at 4 ºC. 60 µL of the lower layer was collected and diluted with 60 µL of $CHCl_3$: MeOH in a glass vial. The samples were kept at -80 ºC until analysis.

The samples were analysed using an ultra-high-performance liquid chromatography quadrupole time-of-flight mass spectrometry (UHPLC-QTOFMS), comprised of a 1290 Infinity UHPLC system coupled to a 6545 QTOF (Agilent Technologies; Santa Clara, CA, USA) in positive ion mode. Samples were randomised prior to analysis. Chromatographic separation was performed using an ACQUITY UPLC BEH C18 column (2.1 mm x 100 mm, particle size 1.7 µm) by Waters (Milford, USA). Quality control was performed throughout the dataset including blanks, pure standard samples, extracted standard samples, QC samples and control plasma samples. The eluent system consisted of (A) 10 mM $NH_4Ac$ in $H_2O$ and 0.1% formic acid and (B) 10 mM $NH_4Ac$ in ACN: IPA (1:1) and 0.1% formic acid. The gradient was as follows: 0–2 min, 35% solvent B; 2–7 min, 80% solvent B; 7–14 min 100% solvent B. The flow rate was 0.4 mL/min. MS data preprocessing was performed using the open-source software MZmine 2.18, using the same workflow as described previously.[2]

A batch correction workflow was performed on the data acquired from both sets using the open-source software QC:MXP (Version 2.0),[3] where the software operates independently and sequentially on each feature based on the assumption that biologically identical QC samples represent accurately the systematic bias (or drift) and random noise in the mass spectrometry measurement process in a batch.[4,5]

Signal drift was observed in the development set, affecting 210 samples (and 5 QCs); these were removed before further analysis, resulting in 1374 samples in the development set, and 559 samples in the test set (unchanged). No comparable drift was seen in the test set. Sample exclusions at this step are shown in **Figure 1**. The remaining samples were reprocessed using QC:MXP (Version 2.0),[3], and filtered with a relative standard deviation (RSD) ≤ 20% and D-ratio ≤ 40% cut-off, retaining 278 variables. Missing lipid values arising from signals below the limit of detection were handled as described under Statistical analysis. Completeness of the clinical covariates is summarised in **Table 1.**

### Statistical analysis

All statistical analyses were performed in R 4.4.3 All preprocessing steps were fitted on the development set and applied unchanged to the test set; no information from the test set contributed to imputation, transformation or scaling parameters. Batch correction is exempt from this concern because QC:MXP operates within each analytical batch using that batch's own pooled QC samples, so no information crosses the development/test split. Missing lipid values, which arise when a feature falls below the limit of detection in a given sample, were imputed with half of the minimum observed value for that feature and the dataset was log-transformed and autoscaled prior to statistical analysis.

Unsupervised lipid clustering was performed using model-based Gaussian finite mixture modelling implemented in the *mclust* package,[6] where the clustering was conducted on the lipid variables to identify groups of lipids with similar profiles across samples. Lipids were selected and assigned to specific clusters based on model selection via Bayesian Information Criterion (BIC), and cluster scores were computed as the mean abundance of all lipids belonging to that cluster. After obtaining the cluster scores, correlation analysis was performed between the lipid clusters, protein biomarker levels available in the CENTER-TBI database (S100B - S100 calcium-binding protein B, NSE - Neuron-specific enolase, GFAP - Glial fibrillary acidic protein, UCH-L1 - Ubiquitin C-terminal hydrolase L1, NFL - Neurofilament light, T-Tau – Total Tau), GOSE outcomes and CRASH covariates (Glasgow Coma Scale at admission, pupillary reactivity, presence or absence of extracranial injuries, defined as Acute Injury Score of 3 and above in any extracranial area, and age).[7] Spearman correlation was performed using the *Hmisc* package, and p-value significances were adjusted using Benjamini-Hochberg false discovery rate (FDR). Correlation plot was built and visualised using the *pheatmap* package, and significant associations were annotated in the plot to facilitate visual interpretation.

To observe if there is any relationship between choline and the outcome of the patients after 6 months, the intensities of all choline-containing lipids (PCs, LPCs and SMs) were summed and visualised in a violin plot for each GOSE level (1, 2/3, 4, 5, 6, 7 and 8), where a Jonckheere-Terpstra test[8,9] was performed to evaluate if there was a monotonic trend across the ordered groups. Violin plots were built using the *ggplot2* package, and the package *DescTools* was used for the Jonckheere-Terpstra test.

Ordinal Logistic Regression was used to evaluate a potential lipid signature that can improve the prediction of TBI outcome at 6 months of injury, considering the ordinality of the GOSE scale, rather than dealing with the outcome scale as several dichotomised endpoints. GOSE scale was collapsed into five ordered levels to ensure sufficient sample sizes (1-2-3-4, death/ vegetative state/ lower severe disability/upper severe disability; 5, lower moderate disability; 6, upper moderate disability; 7, lower good recovery; 8, upper good recovery) to ensure sufficient sample sizes per category, as some individual GOSE levels presented a reduced amount of individuals. Before the ordinal logistic regression workflow, subjects with missing values in any of the covariates (CRASH clinical variables) were excluded using a complete-case approach. Protein and lipid data were complete for all patients. The resulting complete-case dataset in the development set comprised 411 samples, while the held-out test set was comprised of 367 samples. The split was fixed before modelling began and the test set was not inspected at any point during variable selection or model fitting. We deliberately avoid the term external validation, since the test set shares study, biobank, laboratory, storage and analytical platform with the training data, and differs from it only in when it was acquired and in having been assembled as an mTBI subcohort from the outset rather than subsetted from an all-severity sample set.

Modelling workflow consisted of two sequential steps: Variable selection using ordinal elastic-net and proportional odds logistic regression. The primary modelling workflow was performed using a complete-case approach, with no imputation performed before the variable selection or model building. Multiple imputation on covariates data was performed subsequently as a sensitivity analysis to evaluate the impact of missing covariate data on the final model.

Ordinal Elastic-Net was used to identify a set of lipids associated with the ordinal outcome of TBI, using the *ordinalNet* package.[10] Models were fitted using cumulative logit (proportional odds) link function, using $\alpha = 0.5$ to balance Ridge and LASSO penalties. To ensure the robustness of the selected variables, a stability selection scheme was implemented, where the training set was subsampled 20 times, selecting 80% of the observations without replacement, and each subsample was fitted using a 5-fold CV to select the optimal hyperparameter ($\lambda$), using cross-validated log-likelihood as tune method (*cvLoglik*). Variables selected in more than 70% of the stability runs were retained for subsequent analysis.

Proportional odds logistic regression models were built using the *polr* function from the MASS package. The *polr* model is a model built on cumulative *logits*, which calculates the log-odds of being in one side of a cumulative cutoff in a ordered outcome, estimating a single set of predictors effects across all outcomes while allowing threshold specific-intercepts, preserving the natural ordering of the outcome scale, yielding J-1 intercepts for J categories.[11,12] Two models were built using the same individuals as observations: one containing only the CRASH covariates, and other containing the selected lipids, proteins and the CRASH covariates, to evaluate the increment in prediction from the lipid panel across the ordinal outcome scale. Internal validation during model development was performed *via* bootstrapping (2000 training-set iterations), yielding optimism-corrected AUC values in the training set. ROC curves, raw AUC and DeLong 95% confidence intervals were obtained from the held-out test set. AUC performance metrics were evaluated for both models (CRASH covariates only and CRASH covariates plus lipids and proteins). Model discrimination was evaluated through receiver operating characteristic (ROC) curves with the *pROC* package, which were smoothed using a bootstrap-average method (2000 iterations). Area under the curve (AUC) was used as the performance metric, for each threshold of the ordinal outcome. The *polr* ordinal model general performance was also evaluated using the average dichotomous c-index, calculated by computing the average of all J-1 dichotomous thresholds on the test set. AUCs of the two models were compared with DeLong's test at each of the four cumulative GOSE thresholds. We also analysed the added value of the model with lipids and proteins using pseudo-Nagelkerke $R^2$. As a supplementary analysis, ordinal elastic-net variable selection (using the same stability-selection procedure described above) was run separately on the full lipid set and the full protein set, independent of the combined model. In both cases, the variables selected matched those retained by the combined lipid-plus-protein elastic-net run. Using these single-domain variable sets, two additional proportional-odds models were fitted (CRASH plus proteins-only and CRASH plus lipids-only) following the same modelling and validation procedures as the primary analysis, and evaluated using the same AUC, DeLong comparison, average dichotomous c-index, and pseudo-Nagelkerke $R^2$ metrics on the held-out test set. This allowed the discriminative contribution of each biomarker class to be isolated independent of clinical covariates.

Because four comparisons were made, we report unadjusted p-values for comparability with previous TBI biomarker studies but also applied Benjamini–Hochberg correction; results of the unadjusted analysis and analysis with corrections for multiple comparisons are given, and threshold-specific findings are treated as exploratory throughout.

A sensitivity analysis evaluated the robustness of the final ordinal logistic regression model to missing clinical covariate data. Admission GCS and Pupillary reactiveness missing values were imputed by multiple imputation, using the *mice* package. 30 datasets were imputed, using the other covariates and the predictors chosen by Ordinal Elastic-Net. The same ordinal logistic regression was fitted to the imputed datasets, and the estimates were combined using Rubin's rules and compared with the results obtained from complete-case analysis on their respective odds ratio (OR) for each variable.

Annotated lipids that were chosen by the Ordinal Elastic-Net algorithm were visualised as violin plots and evaluated using a Jonckheere-Terpstra test to analyse their possible trends along the outcome scale, the same way as performed with the choline relationship test. Intensities were log-transformed for better visualisation of the violin plots.

### Supplementary Tables

**Table S1.** Cluster assignments for each lipid using the *mclust* algorithm, with their respective *m/z* and retention time.

| Cluster | Lipid Name | m/z | Retention time (min) | Adduct | MSI level |
|---|---|---|---|---|---|
| 1 | CE(18:2) | 369.3515 | 8.85 | Cholesterol fragment | 1 |
| 1 | CE(20:4) | 369.3514 | 8.60 | Cholesterol fragment | 1 |
| 1 | CE(20:5) | 369.3513 | 8.47 | Cholesterol fragment | 2 |
| 1 | LPC(16:0) | 496.3398 | 3.25 | $[M+H]^+$ | 1 |
| 1 | PC(34:3) | 756.5537 | 5.71 | $[M+H]^+$ | 2 |
| 1 | PC(36:3) | 784.5855 | 6.16 | $[M+H]^+$ | 2 |
| 1 | PC(36:3)_2 | 784.5855 | 6.22 | $[M+H]^+$ | 2 |
| 1 | PC(36:4)_2 | 782.5706 | 5.99 | $[M+H]^+$ | 2 |
| 1 | PC(36:4)_3 | 782.5676 | 6.51 | $[M+H]^+$ | 2 |
| 1 | PC(37:4) | 796.5854 | 6.21 | $[M+H]^+$ | 2 |
| 1 | PC(38:4) | 810.6003 | 6.31 | $[M+H]^+$ | 2 |
| 1 | PC(38:4)_2 | 810.6012 | 6.41 | $[M+H]^+$ | 2 |
| 1 | PC(40:4) | 838.6311 | 6.74 | $[M+H]^+$ | 2 |
| 1 | PE(16:0/18:1) | 718.5386 | 6.65 | $[M+H]^+$ | 1 |
| 1 | PE(16:0/20:4) | 740.5223 | 6.11 | $[M+H]^+$ | 2 |
| 1 | PE(18:0/20:4) | 768.5537 | 6.56 | $[M+H]^+$ | 2 |
| 1 | PI(18:0/20:4) | 904.5904 | 6.06 | $[M+H]^+$ | 1 |
| 1 | SM(d18:1/24:0) | 815.6943 | 7.46 | $[M+H]^+$ | 1 |
| 1 | TG(16:0/18:2/18:2) | 872.7707 | 8.42 | $[M+NH4]^+$ | 2 |
| 1 | TG(16:0/18:2/18:2)_2 | 872.7705 | 8.47 | $[M+NH4]^+$ | 2 |
| 1 | TG(16:0/22:5/18:1) or TG(20:4/18:1/18:1) | 924.8016 | 8.47 | $[M+NH4]^+$ | 2 |
| 1 | TG(18:0/18:1/20:4) | 926.8167 | 8.78 | $[M+NH4]^+$ | 2 |
| 1 | TG(18:0/18:1/20:4)_2 | 926.8169 | 8.79 | $[M+NH4]^+$ | 2 |
| 1 | TG(52:4) | 877.7258 | 8.42 | $[M+NH4]^+$ | 2 |
| 1 | TG(52:4)_2 | 877.7256 | 8.46 | $[M+NH4]^+$ | 2 |
| 1 | TG(54:6)_3 | 896.7701 | 8.24 | $[M+NH4]^+$ | 2 |
| 1 | TG(56:6) | 929.7566 | 8.48 | $[M+NH4]^+$ | 2 |
| 1 | Unknown1 | 876.5702 | 6.58 | - | - |
| 1 | Unknown10 | 931.7697 | 8.67 | - | - |
| 1 | Unknown117 | 968.5567 | 6.43 | - | - |
| 1 | Unknown121 | 942.5411 | 6.20 | - | - |
| 1 | Unknown168 | 793.5479 | 6.17 | - | - |
| 1 | Unknown169 | 734.6232 | 6.81 | - | - |
| 1 | Unknown171 | 1159.834 | 6.11 | - | - |
| 1 | Unknown195 | 850.5549 | 6.51 | - | - |
| 1 | Unknown223 | 918.5414 | 6.50 | - | - |
| 1 | Unknown226 | 627.5342 | 6.06 | - | - |

| | | | | | |
|---|---|---|---|---|---|
| 1 | Unknown248 | 944.5569 | 6.58 | - | - |
| 1 | Unknown263 | 909.5458 | 6.06 | - | - |
| 1 | Unknown28 | 900.57 | 6.43 | - | - |
| 1 | Unknown292 | 945.7292 | 8.48 | - | - |
| 1 | Unknown34 | 940.5256 | 5.98 | - | - |
| 1 | Unknown34_2 | 940.5256 | 5.99 | - | - |
| 1 | Unknown45 | 872.539 | 5.99 | - | - |
| 1 | Unknown48 | 774.5635 | 5.60 | - | - |
| 1 | Unknown54 | 1195.834 | 5.99 | - | - |
| 2 | Cer (d40:1) / (d18:1/22:0) | 604.6019 | 7.72 | $[M+H-H2O]^+$ | 2 |
| 2 | Cer(d18:1/22:0) | 622.6127 | 7.72 | $[M+H]^+$ | 1 |
| 2 | Cer(d18:1/23:0) | 636.6284 | 7.88 | $[M+H]^+$ | 2 |
| 2 | Cer(d18:1/24:0) | 650.6443 | 8.03 | $[M+H]^+$ | 1 |
| 2 | Cer(d18:1/24:1) | 648.6282 | 7.65 | $[M+H]^+$ | 1 |
| 2 | Cer(d42:1) | 632.6335 | 8.03 | $[M+H-H2O]^+$ | 2 |
| 2 | PG (O-41:0) | 835.6667 | 7.38 | $[M+H]^+$ | 2 |
| 2 | PI (44:4) | 971.6403 | 7.38 | $[M+H]^+$ | 2 |
| 2 | SM(37:1) | 745.6207 | 6.86 | $[M+H]^+$ | 2 |
| 2 | SM(39:2) | 771.6359 | 6.85 | $[M+H]^+$ | 2 |
| 2 | SM(40:2) | 785.6533 | 7.05 | $[M+H]^+$ | 2 |
| 2 | SM(40:2) Na+ adduct | 807.6348 | 7.06 | $[M+H]^+$ | 2 |
| 2 | SM(d16:1/18:1) or SM(d18:2/16:0) | 701.5593 | 5.62 | $[M+H]^+$ | 2 |
| 2 | SM(d18:1/16:0) | 703.5752 | 6.15 | $[M+H]^+$ | 1 |
| 2 | SM(d18:1/24:0)_2 | 815.7003 | 7.79 | $[M+H]^+$ | 1 |
| 2 | SM(d32:1) | 675.5433 | 5.59 | $[M+H]^+$ | 2 |
| 2 | SM(d33:1) | 689.5588 | 5.88 | $[M+H]^+$ | 2 |
| 2 | SM(d36:1) | 731.6065 | 6.64 | $[M+H]^+$ | 2 |
| 2 | SM(d36:2) | 729.5904 | 6.16 | $[M+H]^+$ | 2 |
| 2 | SM(d38:2) | 757.6217 | 6.63 | $[M+H]^+$ | 2 |
| 2 | SM(d39:1) | 773.6527 | 7.26 | $[M+H]^+$ | 2 |
| 2 | SM(d40:1) | 787.6692 | 7.45 | $[M+H]^+$ | 2 |
| 2 | SM(d41:1) | 801.6844 | 7.63 | $[M+H]^+$ | 2 |
| 2 | SM(d41:2) | 799.6686 | 7.23 | $[M+H]^+$ | 2 |
| 2 | SM(d42:2) | 813.6849 | 7.38 | $[M+H]^+$ | 2 |
| 2 | SM(d42:3) | 811.6691 | 6.98 | $[M+H]^+$ | 2 |
| 2 | TG(48:4) | 821.6499 | 7.23 | $[M+NH4]^+$ | 2 |
| 2 | Unknown103 | 903.6532 | 7.38 | - | - |
| 2 | Unknown119 | 630.6174 | 7.65 | - | - |
| 2 | Unknown140 | 715.5739 | 5.90 | - | - |
| 2 | Unknown147 | 827.6992 | 7.49 | - | - |
| 2 | Unknown151 | 825.6823 | 7.17 | - | - |
| 2 | Unknown157 | 945.6246 | 7.45 | - | - |
| 2 | Unknown161 | 877.6375 | 7.45 | - | - |
| 2 | Unknown189 | 831.6937 | 7.18 | - | - |
| 2 | Unknown24 | 797.6489 | 6.79 | - | - |
| 2 | Unknown267 | 841.7129 | 7.74 | - | - |

| | | | | | |
|---|---|---|---|---|---|
| 2 | Unknown59 | 1017.935 | 8.84 | - | - |
| 2 | Unknown6 | 784.6382 | 6.58 | - | - |
| 2 | Unknown71 | 901.6373 | 6.99 | - | - |
| 3 | HexCer(d18:1/24:0) | 812.6962 | 7.77 | $[M+H]^+$ | 1 |
| 3 | PC (O-42:5) /(o-22:1/20:4) | 850.6671 | 7.01 | $[M+H]^+$ | 2 |
| 3 | PC (36:4) /(18:3/18:1) | 788.5568 | 6.18 | $[M+H]^+$ | 2 |
| 3 | PC (39:0) / (13:0/26:0) | 832.6631 | 7.39 | $[M+H]^+$ | 2 |
| 3 | PC(16:0e/18:1(9Z)) | 746.6062 | 6.79 | $[M+H]^+$ | 1 |
| 3 | PC(18:0p/18:1(9Z)) | 772.622 | 6.81 | $[M+H]^+$ | 1 |
| 3 | PC(O-32:0) | 720.5896 | 6.81 | $[M+H]^+$ | 2 |
| 3 | PC(O-32:1) | 718.574 | 6.74 | $[M+H]^+$ | 2 |
| 3 | PC(O-34:2) | 744.5899 | 6.41 | $[M+H]^+$ | 2 |
| 3 | PC(O-34:3) | 742.5748 | 6.33 | $[M+H]^+$ | 2 |
| 3 | PC(O-36:3) | 770.603 | 6.37 | $[M+H]^+$ | 2 |
| 3 | PC(O-36:3)_3 | 770.6034 | 6.39 | $[M+H]^+$ | 2 |
| 3 | PC(O-36:3)_2 | 770.604 | 6.75 | $[M+H]^+$ | 2 |
| 3 | PC(O-36:4) | 768.5901 | 6.29 | $[M+H]^+$ | 2 |
| 3 | PC(O-36:4)_2 | 768.5897 | 6.31 | $[M+H]^+$ | 2 |
| 3 | PC(O-36:5) | 766.5744 | 6.18 | $[M+H]^+$ | 2 |
| 3 | PC(O-38:4) | 796.6202 | 6.66 | $[M+H]^+$ | 2 |
| 3 | PC(O-38:4)_4 | 796.6208 | 6.70 | $[M+H]^+$ | 2 |
| 3 | PC(O-38:4)_3 | 796.6182 | 6.50 | $[M+H]^+$ | 2 |
| 3 | PC(O-38:4)_2 | 796.6202 | 6.61 | $[M+H]^+$ | 2 |
| 3 | PC(O-38:5) | 794.6058 | 6.25 | $[M+H]^+$ | 2 |
| 3 | PC(O-38:5)_2 | 794.6047 | 6.61 | $[M+H]^+$ | 2 |
| 3 | PC(O-38:6)_2 | 792.5895 | 6.10 | $[M+H]^+$ | 2 |
| 3 | PC(O-38:6)_4 | 792.5895 | 6.13 | $[M+H]^+$ | 2 |
| 3 | PC(O-38:6)_3 | 792.5896 | 6.12 | $[M+H]^+$ | 2 |
| 3 | PC(O-40:5) | 822.6356 | 6.65 | $[M+H]^+$ | 2 |
| 3 | PC(O-40:5)_2 | 822.6356 | 6.66 | $[M+H]^+$ | 2 |
| 3 | PC(O-40:6) | 820.6189 | 6.23 | $[M+H]^+$ | 2 |
| 3 | PC(O-44:5) | 878.6989 | 7.34 | $[M+H]^+$ | 2 |
| 3 | PE(16:1e/20:3) | 726.5433 | 6.43 | $[M+H]^+$ | 2 |
| 3 | PE(O-38:5) or PE(P-38:4) | 752.557 | 6.75 | $[M+H]^+$ | 2 |
| 3 | PE(P-18:0/18:2) | 728.5581 | 6.89 | $[M+H]^+$ | 2 |
| 3 | SM(d18:0/16:0) | 705.5875 | 6.29 | $[M+H]^+$ | 2 |
| 3 | Unknown111 | 806.6473 | 7.46 | - | - |
| 3 | Unknown155 | 834.6786 | 7.77 | - | - |
| 3 | Unknown162 | 824.6513 | 7.07 | - | - |
| 3 | Unknown17 | 772.6205 | 7.10 | - | - |
| 3 | Unknown21 | 876.6827 | 6.99 | - | - |
| 3 | Unknown57 | 854.697 | 7.45 | - | - |
| 3 | Unknown64 | 774.6373 | 7.17 | - | - |
| 3 | Unknown73 | 848.6504 | 6.67 | - | - |
| 3 | Unknown89 | 852.6801 | 7.40 | - | - |
| 3 | Unknown9 | 744.5886 | 6.71 | - | - |

| | | | | | |
|---|---|---|---|---|---|
| 3 | Unknown98 | 722.5542 | 6.24 | - | - |
| 4 | LPC(14:0) | 468.3076 | 2.88 | $[M+H]^+$ | 1 |
| 4 | LPC(16:0e) | 482.3609 | 3.48 | $[M+H]^+$ | 2 |
| 4 | LPC(18:0) | 524.3712 | 3.73 | $[M+H]^+$ | 1 |
| 4 | LPC(18:1) | 522.3554 | 3.32 | $[M+H]^+$ | 1 |
| 4 | LPC(18:2) | 520.3395 | 3.04 | $[M+H]^+$ | 1 |
| 4 | LPC(20:4) | 544.3392 | 3.00 | $[M+H]^+$ | 1 |
| 4 | LPC(20:5) | 542.3212 | 3.04 | $[M+H]^+$ | 1 |
| 4 | Unknown69 | 508.3759 | 3.52 | - | - |
| 4 | Unknown90 | 482.3233 | 3.07 | - | - |
| 5 | PC (38:2) / (14:1/24:1) | 814.6289 | 6.81 | $[M+H]^+$ | 2 |
| 5 | PC(16:0/16:0) | 734.5699 | 6.54 | $[M+H]^+$ | 1 |
| 5 | PC(16:0/18:1) | 760.5864 | 6.52 | $[M+H]^+$ | 1 |
| 5 | PC(30:0) | 706.5379 | 6.05 | $[M+H]^+$ | 2 |
| 5 | PC(32:1) | 732.5539 | 6.06 | $[M+H]^+$ | 2 |
| 5 | PC(32:2) | 730.5377 | 5.62 | $[M+H]^+$ | 2 |
| 5 | PC(33:0) | 748.5837 | 6.67 | $[M+H]^+$ | 2 |
| 5 | PC(34:2) | 758.571 | 6.12 | $[M+H]^+$ | 2 |
| 5 | PC(35:4) | 768.5529 | 5.74 | $[M+H]^+$ | 2 |
| 5 | PC(36:1) | 788.6168 | 6.92 | $[M+H]^+$ | 2 |
| 5 | PC(36:2) | 744.5534 | 5.88 | $[M+H]^+$ | 2 |
| 5 | PC(36:2)_2 | 786.6016 | 6.56 | $[M+H]^+$ | 2 |
| 5 | PC(36:3)_3 | 784.5856 | 6.15 | $[M+H]^+$ | 2 |
| 5 | PC(36:4) | 782.5688 | 5.73 | $[M+H]^+$ | 2 |
| 5 | PC(37:2) | 800.6161 | 6.76 | $[M+H]^+$ | 2 |
| 5 | PC(37:3) | 798.5997 | 6.35 | $[M+H]^+$ | 2 |
| 5 | PC(37:3)_2 | 798.6002 | 6.38 | $[M+H]^+$ | 2 |
| 5 | PC(37:3)_3 | 798.6007 | 6.41 | $[M+H]^+$ | 2 |
| 5 | PC(38:3) | 812.6168 | 6.63 | $[M+H]^+$ | 2 |
| 5 | PC(40:5) | 836.6162 | 6.39 | $[M+H]^+$ | 2 |
| 5 | PC(40:8) | 830.5684 | 5.40 | $[M+H]^+$ | 2 |
| 5 | PE(18:1/18:2) | 742.5355 | 6.19 | $[M+H]^+$ | 2 |
| 5 | PS(41:5)_2 | 874.5544 | 6.20 | $[M+H]^+$ | 2 |
| 5 | PS(41:6) | 850.5685 | 6.62 | $[M+H]^+$ | 2 |
| 5 | SM(d18:1/12:0) | 647.5109 | 5.00 | $[M+H]^+$ | 2 |
| 5 | Unknown139 | 704.5248 | 5.56 | - | - |
| 5 | Unknown31 | 754.5365 | 5.49 | - | - |
| 5 | Unknown33 | 878.5854 | 6.92 | - | - |
| 5 | Unknown35 | 946.5724 | 6.92 | - | - |
| 5 | Unknown58 | 902.5848 | 6.62 | - | - |
| 5 | Unknown79 | 970.5721 | 6.63 | - | - |
| 5 | Unknown81 | 802.6315 | 7.11 | - | - |
| 6 | LPC(22:6) | 568.339 | 2.94 | $[M+H]^+$ | 1 |
| 6 | PC (38:1) | 816.6468 | 7.27 | $[M+H]^+$ | 2 |
| 6 | PC(18:0p/22:6) | 818.6043 | 6.41 | $[M+H]^+$ | 1 |
| 6 | PC(36:5) | 780.5541 | 5.63 | $[M+H]^+$ | 2 |

| | | | | | |
|---|---|---|---|---|---|
| 6 | PC(37:5) | 794.5688 | 5.80 | $[M+H]^+$ | 2 |
| 6 | PC(38:6) | 806.57 | 5.80 | $[M+H]^+$ | 2 |
| 6 | PC(40:6) | 834.6013 | 6.25 | $[M+H]^+$ | 2 |
| 6 | PC(40:7) | 832.5843 | 5.79 | $[M+H]^+$ | 2 |
| 6 | PC(O-36:5)_2 | 766.5731 | 6.00 | $[M+H]^+$ | 2 |
| 6 | PC(O-38:6) | 792.5886 | 5.95 | $[M+H]^+$ | 2 |
| 6 | PC(O-40:6)_2 | 820.6206 | 6.50 | $[M+H]^+$ | 2 |
| 6 | PC(P-18:0/22:6) | 818.6054 | 6.05 | $[M+H]^+$ | 1 |
| 6 | PE(18:1e/22:6) | 776.5596 | 6.56 | $[M+H]^+$ | 2 |
| 6 | PE(P-16:0/22:6) | 748.5269 | 6.14 | $[M+H]^+$ | 2 |
| 6 | Unknown129 | 964.5254 | 5.80 | - | - |
| 6 | Unknown137 | 924.5693 | 6.25 | - | - |
| 6 | Unknown144 | 992.5563 | 6.25 | - | - |
| 6 | Unknown154 | 896.5386 | 5.80 | - | - |
| 6 | Unknown175 | 874.6671 | 6.84 | - | - |
| 6 | Unknown53 | 764.5582 | 5.83 | - | - |
| 6 | Unknown76 | 902.6985 | 7.17 | - | - |
| 6 | Unknown93 | 792.553 | 5.56 | - | - |
| 7 | TG(18:1/18:1/16:0) | 876.8019 | 9.00 | $[M+NH4]^+$ | 2 |
| 7 | TG(18:1/18:1/18:1) | 902.8174 | 8.93 | $[M+NH4]^+$ | 2 |
| 7 | TG(18:1/18:2/18:2) | 898.7862 | 8.38 | $[M+NH4]^+$ | 2 |
| 7 | TG(18:2/18:1/16:0) | 874.7866 | 8.68 | $[M+NH4]^+$ | 2 |
| 7 | TG(18:2/18:1/18:1) | 900.802 | 8.63 | $[M+NH4]^+$ | 2 |
| 7 | TG(50:1) | 855.7415 | 8.44 | $[M+NH4]^+$ | 2 |
| 7 | TG(51:4) | 858.7535 | 8.29 | $[M+NH4]^+$ | 2 |
| 7 | TG(52:2) | 881.7573 | 9.00 | $[M+NH4]^+$ | 2 |
| 7 | TG(52:3) | 879.7419 | 8.68 | $[M+NH4]^+$ | 2 |
| 7 | TG(53:2) | 890.8166 | 9.18 | $[M+NH4]^+$ | 2 |
| 7 | TG(53:3) | 888.801 | 8.82 | $[M+NH4]^+$ | 2 |
| 7 | TG(53:3)_2 | 893.7563 | 8.82 | $[M+NH4]^+$ | 2 |
| 7 | TG(53:4) | 886.7849 | 8.51 | $[M+NH4]^+$ | 2 |
| 7 | TG(54:3) | 907.7726 | 8.93 | $[M+NH4]^+$ | 2 |
| 7 | TG(54:4) | 905.7572 | 8.63 | $[M+NH4]^+$ | 2 |
| 7 | TG(54:5) | 903.7408 | 8.38 | $[M+NH4]^+$ | 2 |
| 7 | TG(56:4) | 928.8293 | 8.86 | $[M+NH4]^+$ | 2 |
| 7 | Unknown210 | 577.5186 | 9.00 | - | - |
| 7 | Unknown307 | 925.7608 | 9.33 | - | - |
| 7 | Unknown32 | 859.7736 | 9.00 | - | - |
| 7 | Unknown87 | 895.7715 | 9.16 | - | - |
| 8 | TG(14:0/16:0/18:1) | 822.7548 | 8.77 | $[M+NH4]^+$ | 2 |
| 8 | TG(14:0/18:1/18:1) | 848.7708 | 8.73 | $[M+NH4]^+$ | 2 |
| 8 | TG(14:0/18:2/18:2) | 844.7385 | 8.17 | $[M+NH4]^+$ | 2 |
| 8 | TG(16:0/16:0/12:0) | 768.7068 | 8.55 | $[M+NH4]^+$ | 2 |
| 8 | TG(16:1/18:1/12:0) | 794.7229 | 8.49 | $[M+NH4]^+$ | 2 |
| 8 | TG(18:1/12:0/18:1) or TG(18:2/16:0/14:0) | 820.739 | 8.46 | $[M+NH4]^+$ | 2 |
| 8 | TG(46:0) | 796.7383 | 8.84 | $[M+NH4]^+$ | 2 |

| | | | | | |
|---|---|---|---|---|---|
| 8 | TG(46:2) | 792.707 | 8.20 | $[M+NH4]^+$ | 2 |
| 8 | TG(48:1) | 827.7096 | 8.78 | $[M+NH4]^+$ | 2 |
| 8 | TG(48:2) | 825.6937 | 8.46 | $[M+NH4]^+$ | 2 |
| 8 | TG(48:3) | 818.7226 | 8.17 | $[M+NH4]^+$ | 2 |
| 8 | TG(49:1) | 836.7695 | 8.92 | $[M+NH4]^+$ | 2 |
| 8 | TG(49:2) | 834.7538 | 8.58 | $[M+NH4]^+$ | 2 |
| 8 | TG(50:1)_2 | 850.7861 | 9.07 | $[M+NH4]^+$ | 2 |
| 8 | TG(50:2) | 853.7259 | 8.74 | $[M+NH4]^+$ | 2 |
| 8 | TG(50:3) | 846.755 | 8.42 | $[M+NH4]^+$ | 2 |
| 8 | TG(50:5) | 842.721 | 7.97 | $[M+NH4]^+$ | 2 |
| 8 | TG(51:2) | 862.7852 | 8.84 | $[M+NH4]^+$ | 2 |
| 8 | TG(51:2)_2 | 862.7849 | 9.24 | $[M+NH4]^+$ | 2 |
| 8 | Unknown12 | 883.7724 | 9.41 | - | - |
| 8 | Unknown13 | 855.7413 | 9.07 | - | - |
| 8 | Unknown14 | 605.5491 | 9.36 | - | - |
| 8 | Unknown14_2 | 605.5492 | 9.38 | - | - |
| 8 | Unknown23 | 766.6913 | 8.23 | - | - |
| 8 | Unknown258 | 764.6754 | 7.97 | - | - |
| 8 | Unknown295 | 899.7453 | 9.43 | - | - |
| 8 | Unknown30 | 551.5025 | 8.75 | - | - |
| 8 | Unknown60 | 575.5026 | 8.68 | - | - |
| 8 | Unknown60_2 | 575.5027 | 8.69 | - | - |
| 8 | Unknown74 | 551.5029 | 9.08 | - | - |
| 8 | Unknown74_2 | 551.503 | 9.09 | - | - |
| 8 | Unknown78 | 549.4869 | 8.57 | - | - |
| 8 | Unknown97 | 849.6935 | 8.17 | - | - |
| 9 | PE(16:0/22:6) | 764.5221 | 5.93 | $[M+H]^+$ | 1 |
| 9 | TG(16:0/18:2/22:6) | 920.7697 | 8.07 | $[M+NH4]^+$ | 2 |
| 9 | TG(16:0/18:2/22:6)_2 | 920.77 | 8.11 | $[M+NH4]^+$ | 2 |
| 9 | TG(16:0/22:5/18:1) or TG(20:4/18:1/18:1)_2 | 924.8018 | 8.48 | $[M+NH4]^+$ | 2 |
| 9 | TG(18:0/18:1/20:4)_3 | 926.8171 | 8.80 | $[M+NH4]^+$ | 2 |
| 9 | TG(18:1/18:1/22:6) | 948.8011 | 8.32 | $[M+NH4]^+$ | 2 |
| 9 | TG(18:1/18:2/18:2)_2 | 898.7862 | 8.52 | $[M+NH4]^+$ | 2 |
| 9 | TG(18:2/18:2/18:2) or TG(18:3/18:2/18:1) | 896.7702 | 8.20 | $[M+NH4]^+$ | 2 |
| 9 | TG(18:2/22:5/16:0) | 922.7861 | 8.27 | $[M+NH4]^+$ | 2 |
| 9 | TG(18:2/22:5/16:0)_2 | 922.7862 | 8.32 | $[M+NH4]^+$ | 2 |
| 9 | TG(54:5)_2 | 903.741 | 8.51 | $[M+NH4]^+$ | 2 |
| 9 | TG(54:6) | 901.725 | 8.20 | $[M+NH4]^+$ | 2 |
| 9 | TG(54:6)_4 | 901.7251 | 8.20 | $[M+NH4]^+$ | 2 |
| 9 | TG(54:6)_2 | 901.7251 | 8.20 | $[M+NH4]^+$ | 2 |
| 9 | TG(54:7) | 894.7538 | 7.99 | $[M+NH4]^+$ | 2 |
| 9 | TG(54:7)_2 | 894.754 | 8.08 | $[M+NH4]^+$ | 2 |
| 9 | TG(56:6)_2 | 929.7564 | 8.47 | $[M+NH4]^+$ | 2 |
| 9 | TG(56:9) | 918.7534 | 7.87 | $[M+NH4]^+$ | 2 |
| 9 | TG(58:10) | 944.769 | 7.89 | $[M+NH4]^+$ | 2 |
| 9 | TG(58:10)_2 | 944.769 | 7.91 | $[M+NH4]^+$ | 2 |

| | | | | | |
|---|---|---|---|---|---|
| 9 | TG(58:6) | 952.8303 | 8.66 | $[M+NH4]^+$ | 2 |
| 9 | TG(58:6)_2 | 952.8308 | 8.68 | $[M+NH4]^+$ | 2 |
| 9 | TG(58:6)_3 | 952.8303 | 8.66 | $[M+NH4]^+$ | 2 |
| 9 | TG(58:9)_2 | 946.7846 | 8.08 | $[M+NH4]^+$ | 2 |
| 9 | Unknown107 | 951.7403 | 8.09 | - | - |
| 9 | Unknown114 | 925.7249 | 8.08 | - | - |
| 9 | Unknown114_2 | 925.7251 | 8.10 | - | - |
| 9 | Unknown159 | 899.7092 | 8.04 | - | - |
| 9 | Unknown159_2 | 899.7092 | 8.07 | - | - |
| 9 | Unknown182 | 927.7409 | 8.25 | - | - |
| 9 | Unknown199 | 970.7847 | 7.96 | - | - |

**Table S2.** Optimism-corrected AUCs (training set) for the baseline model and for the baseline with proteins and lipids added, AUC values for the baseline model (test set), the AUC for the baseline with proteins and lipids added (test set), Delta AUC, unadjusted p-value (Delong) and adjusted p-values (Benjamini-Hochberg) for each individual GOSE threshold.

| Threshold | Baseline Optimism-corrected AUC | Baseline + proteins + lipids Optimism-corrected AUC | Baseline AUC (Test set) | Baseline + proteins + lipids AUC (Test set) | Δ AUC (Test set) | p-value (Delong) | p-value (Benjamini-Hochberg) |
|---|---|---|---|---|---|---|---|
| GOSE ≥ 5 | 0.772 | 0.869 | 0.797 | 0.802 | 0.005 | 0.874 | 0.874 |
| GOSE ≥ 6 | 0.725 | 0.797 | 0.749 | 0.770 | 0.021 | 0.456 | 0.608 |
| GOSE ≥ 7 | 0.669 | 0.732 | 0.673 | 0.728 | 0.055 | 0.022 | 0.088 |
| GOSE = 8 | 0.661 | 0.681 | 0.661 | 0.702 | 0.041 | 0.0748 | 0.150 |

**Table S3.** AUC values for the baseline model (test set), AUC for the baseline with proteins and lipids added individually (test set), Delta AUC, unadjusted p-value (Delong), adjusted p-values (Benjamini-Hochberg), Nagelkerke $R^2$ and dichotomous ordinal c-index with confidence intervals (CI) for supplementary models containing CRASH plus proteins only and CRASH plus lipids only.

**CRASH + Proteins only**

| Threshold | Baseline AUC (Test set) | Baseline + proteins AUC (Test set) | Δ AUC | p-value (Delong) | p-value (Benjamini-Hochberg) |
|---|---|---|---|---|---|
| **GOSE ≥ 5** | 0.797 | 0.82 | 0.023 | 0.358 | 0.358 |
| **GOSE ≥ 6** | 0.749 | 0.773 | 0.024 | 0.293 | 0.358 |
| **GOSE ≥ 7** | 0.673 | 0.708 | 0.035 | 0.0747 | 0.213 |
| **GOSE = 8** | 0.661 | 0.691 | 0.031 | 0.107 | 0.213 |

| Model | Nagelkerke $R^2$ | Δ $R^2$ |
|---|---|---|
| **Baseline** | 0.125 | |
| **Baseline + Proteins** | 0.184 | 0.059 |

| Ordinal C-Index (ORC) | | |
|---|---|---|
| **ORC** | **Lower CI** | **Higher CI** |
| 0.748 | 0.697 | 0.794 |

**CRASH + Lipids only**

| Threshold | Baseline AUC (Test set) | Baseline + lipids AUC (Test set) | Δ AUC | p-value (Delong) | p-value (Benjamini-Hochberg) |
|---|---|---|---|---|---|
| **GOSE ≥ 5** | 0.797 | 0.777 | -0.021 | 0.494 | 0.659 |
| **GOSE ≥ 6** | 0.749 | 0.745 | -0.004 | 0.885 | 0.885 |
| **GOSE ≥ 7** | 0.673 | 0.716 | 0.043 | 0.0704 | 0.281 |
| **GOSE = 8** | 0.661 | 0.689 | 0.028 | 0.194 | 0.388 |

| Model | Nagelkerke $R^2$ | Δ $R^2$ |
|---|---|---|
| **Baseline** | 0.125 | |
| **Baseline + Lipids** | 0.216 | 0.092 |

| Ordinal C-Index (ORC) | | |
|---|---|---|
| **ORC** | **Lower CI** | **Higher CI** |
| 0.732 | 0.681 | 0.781 |

**Table S4.** Sensitivity analysis comparing complete cases and multiple imputed (MI) proportional odds logistic regression models

| Variable | Complete-case OR (95% CI) | *p* | MI OR (95% CI) | *p* |
|---|---|---|---|---|
| S100B | 0.74 (0.58–0.95) | 0.018 | 0.70 (0.56–0.88) | 0.002 |
| GFAP | 0.94 (0.69–1.29) | 0.701 | 1.00 (0.76–1.33) | 0.985 |
| NFL | 0.85 (0.61–1.20) | 0.356 | 0.79 (0.59–1.06) | 0.119 |
| LPC(18:2) | 0.82 (0.25–2.69) | 0.741 | 0.73 (0.25–2.15) | 0.568 |
| LPC(20:5) | 1.31 (0.41–4.27) | 0.648 | 1.39 (0.48–4.02) | 0.547 |
| PC(O-34:3) | 1.08 (0.83–1.42) | 0.561 | 1.13 (0.89–1.44) | 0.329 |
| Unknown1 | 1.31 (1.06–1.61) | 0.011 | 1.23 (1.02–1.49) | 0.028 |
| Unknown90 | 1.15 (0.90–1.48) | 0.263 | 1.19 (0.95–1.49) | 0.137 |
| Unknown69 | 1.10 (0.86–1.41) | 0.428 | 1.13 (0.91–1.40) | 0.287 |
| Unknown157 | 1.23 (0.94–1.61) | 0.123 | 1.12 (0.90–1.39) | 0.317 |
| Age | 0.99 (0.98–1.00) | 0.085 | 0.99 (0.98–1.00) | 0.031 |
| Admission GCS | 1.16 (0.95–1.42) | 0.137 | 1.22 (0.99–1.51) | 0.059 |
| Pupillary reactivity | 1.06 (0.27–4.23) | 0.934 | 0.66 (0.21–2.00) | 0.458 |
| Extracranial injury | 0.60 (0.37–0.98) | 0.041 | 0.64 (0.41–0.97) | 0.038 |
